# Comparative performance of between-population vaccine allocation strategies with applications for emerging pandemics

**DOI:** 10.1101/2021.06.18.21259137

**Authors:** Keya Joshi, Eva Rumpler, Lee Kennedy-Shaffer, Rafia Bosan, Marc Lipsitch

**Author notes:** Contributed equally to this work (Keya Joshi, Eva Rumpler).

## Abstract

Vaccine allocation decisions during emerging pandemics have proven to be challenging due to competing ethical, practical, and political considerations. Complicating decision making, policy makers need to consider vaccine allocation strategies that balance needs both within and between populations. Due to limited vaccine stockpiles, vaccine doses should be allocated in locations where their impact will be maximized. Using a susceptible-exposed-infectious-recovered (SEIR) model we examine optimal vaccine allocation decisions across two populations considering the impact of population size, underlying immunity, continuous vaccine roll-out, heterogeneous population risk structure, and differences in disease transmissibility. We find that in the context of an emerging pathogen where many epidemiologic characteristics might not be known, equal vaccine allocation between populations performs optimally in most scenarios. In the specific case considering heterogeneous population risk structure, first targeting individuals at higher risk of transmission or death due to infection leads to equal resource allocation across populations.

## 3 Introduction

In the past two decades, the 2009 H1N1 influenza pandemic and the recent 2019 SARS-CoV-2 pandemic have highlighted the need for control and mitigation measures against emerging pathogens. SARS-CoV-2 has caused considerable morbidity and mortality, resulting in over 542 million cases and 6.3 million deaths worldwide as of June 2022 (Dong et al., 2020). Previously, the 2009 Swine flu pandemic was estimated to have caused around 200 thousand deaths in the first twelve months globally (Dawood et al., 2012; Simonsen et al., 2013). Vaccines are currently the most effective public health intervention available against emerging pathogens, and have greatly reduced severe disease outcomes (Watson et al., 2022).

Even with the approval and availability of vaccines, both the H1N1 and SARS-CoV-2 pandemics have highlighted key challenges in in the roll-out and uptake of vaccines globally during an ongoing epidemic. During the 2009 H1N1 influenza pandemic, global influenza A vaccine supply was much lower than initially estimated, resulting in large inequities in vaccine access across countries (Fidler, 2010; Kaiser Family Foundation, 2009). In the aftermath of the 2009 H1N1 influenza pandemic the World health Organization (WHO) developed a preparedness framework for the sharing of vaccines in a timely manner, and encouraged advance agreements for vaccine allocation and delivery to improve pandemic response globally (Fineberg, 2014; World Health Organization, 2011).

The COVID-19 pandemic has seen similar challenges, with vaccine supply falling far short of demand. Even with the approval of multiple vaccines, distributed across different regions globally, roll-out has been slow, with vaccination rates highly unequal across countries (Mathieu et al., 2021; Rydland et al., 2022). At the end of 2021, some countries had already vaccinated over 90% of their population, while others did not have access to vaccines (Mathieu et al., 2021). Overall, at the beginning of emerging pandemics, vaccination allocation decisions have been made under the constraint of a limited vaccine stockpile and multiple factors need to be considered to maximize the effect of each dose both within and across populations.

Across both pandemics, numerous papers have shown targeting specific subgroups within the population can result in decreased disease-related morbidity and mortality. For 2009 H1N1 influenza, models found prioritizing individuals at highest risk of complications resulted in the lowest morbidity and mortality (Chowell et al., 2009; Lee et al., 2010; Tuite et al., 2010). For the 2019 SARS-

CoV-2 pandemic, previous work (Bubar et al., 2021; Hogan et al., 2020; Matrajt et al., 2021) has shown, targeting specific subgroups within a given population, including older individuals, results in decreased COVID-19 morbidity and mortality. Other papers looking at health and occupational risk factors (Buckner et al., 2021; Islam et al., 2021) found prioritizing certain occupational groups including healthcare workers, and other essential workers also decreased COVID-19 morbidity and mortality. However, previous theoretical work (Keeling & Shattock, 2012) has also shown that unequal vaccine allocations might be favorable in emerging infectious disease settings, but are less optimal when incorporating realistic assumptions about population heterogeneity and contact structure. This leaves a potentially conflicting message for policy makers when considering optimal allocation strategies. We build upon this work, by not only considering the optimal decision, but also how the decision compares to all possible allocations across two populations. Illustrating these tradeoffs with a simplified model of an emerging pathogen similar to H1N1 or SARS-CoV-2, our results show that the efficiency gains for unequal allocations that are found in models with highly simplified epidemics are typically small; moreover, they vanish and can even reverse under settings more relevant to pandemics caused by emerging pathogens. Similar to previous findings, we show in more realistic scenarios, incorporating population heterogeneity and interaction between populations, that equal distributions are not only optimal, but vastly outperform unequal distributions.

## 4 Materials & Methods

We use a deterministic, two-population, susceptible-exposed-infectious-recovered (SEIR) compartmental model. We assume people are initially susceptible (S). Susceptible individuals move to the exposed state (E) after an effective contact with an infectious individual. After a latent period, exposed individuals become infectious (I). After the infectious period has elapsed, infectious individuals move to a recovered state (R). We do not account for waning immunity and assume once individuals have recovered, they stay immune to infection for the duration of our simulation, here modeled as three years. We start by assuming that there is no interaction between the two populations, so all disease transitions happen in parallel between the two populations.

We extend this SEIR model to allow for underlying immunity (Figure 3, S9) and vaccination (all Figures). At the start of the epidemic, in each population, individuals can be in the susceptible (S), infectious (I), or recovered (R) compartments. When there is underlying immunity, a set proportion of individuals are placed in R. Individuals in R, whether through underlying immunity or infection through the course of the simulation, can never be re-infected. When vaccine doses are distributed to the population, vaccinated individuals are placed in R if the vaccination is successful. We assume that the vaccine is all-or-nothing with 95% efficacy, meaning 95% of those who are vaccinated are placed in R and the remainder stay in S. When there is underlying immunity and vaccination, immune individuals may be vaccinated; vaccination has no effect on them, and they remain in R. Finally, we initialize each simulation by placing 0.1% of individuals in I and allow the epidemic to run, unmitigated except by vaccination, through each population. Full model parameters and equations are shown in SI Appendix A.2. Where possible, parameters represent estimates from both the 2009 H1N1 influenza pandemic and the 2019 SARS-CoV-2 pandemic; for example, 95% vaccine efficacy is close to that estimated for the Moderna mRNA vaccine Spikevax (Doria-Rose et al., 2021).

To recreate the results of Keeling and Shattock (Keeling & Shattock, 2012) we model two homogeneous populations with identical characteristics apart from population size. In this scenario we assume population 2 is double the size of population 1 (Figure 2). For later scenarios, which consider the impact of heterogeneity within populations, we simulate two populations that are identical in size, but vary in their population characteristics (e.g., fraction high risk) (Figure 5, S3-S7).

Next, we extend the model by allowing for heterogeneous risk groups. First, within each population, we model efficient transmitters of infection, for example young adults or children (Goldstein et al., 2021; Kissler et al., 2020) (Figure 5, S3, S4, S6). In this scenario, we assume high-transmitters are four times more likely to transmit compared to low transmitters (Monod et al., 2021). We fix the within-population structure to allow the global *R*_0_ to equal 2, 4, 8 or 16. The full derivation is shown in SI Appendix A.2.6. The SEIR model equation are in SI Appendix A.2.4.

Second, we instead model individuals at elevated risk of death from infection (Figure 5, S3, S5, S7). For COVID-19 this can represent, for example, elderly individuals or other individuals with co-morbidities known to exacerbate disease (C. Wu et al., 2020; Zhou et al., 2020). For the 2009 H1N1 influenza pandemic, this represents young adults. For simplicity of the model, we assume these individuals are five times more likely to die than other infected individuals (Lee et al., 2010; Presanis et al., 2009; Williamson et al., 2020). Note that infection fatality rates are assumed to be constant throughout the epidemic, which may not be realistic as health care resources are strained by large caseloads or case management improves over time. The SEIR model equations are in SI Appendix A.2.5.

Similar to the homogeneous two-population scenario described above, we initialize the model by placing individuals from each population in the susceptible or recovered state based on the pre-existing immunity level and the vaccine allocation scenario. The total number of vaccine doses are split amongst the two populations based on the scenario. Within each population, high-risk individuals are vaccinated first, with leftover doses then allocated to the low-risk population, as described in SI Appendix A.2.7.

Finally, we model the scenario where vaccines are unavailable at the start of the epidemic but are progressively rolled out over the course of the epidemic (Figure 4, S4-S7). For this simulation we vary both the timing of roll-out, and the fraction of the population vaccinated each day. We allow vaccine roll-out to start 1, 10, 30, 50, or 100 days after the epidemic has begun and vary the proportion of the population vaccinated from 1% to 3% per day. For these simulations, the vaccine is allocated within and across populations identically to the scenarios described above for the homogeneous and heterogeneous scenarios.

For each simulation we calculate the cumulative number of infections and deaths from the deterministic SEIR model at the end of the epidemic. Across each scenario we define the optimal allocation strategy as the one that minimizes the total epidemic size (cumulative number of infections) across both populations. Within the high morbidity scenario, we define the optimal allocation strategy as the one that minimizes the total number of deaths across both populations. This is equivalent to maximizing the total number of people across both populations that escape infection (or death) (Duijzer et al., 2018).

We conduct sensitivity analyses to assess the robustness of our results. First, we model a leaky vaccine scenario where we assume the vaccine reduces susceptibility to infection for each individual by 95% (Figure S1). As a result, all vaccinated individuals (except those previously immune through natural infection) can become infected with the virus, although the probability of infection for each contact with an infected individual is lower than for an unvaccinated individual. The SEIR model equations are in SI Appendix A.2.3. We further extend the model by relaxing the assumption that the two populations do not interact (Figure S2). We allow a fraction *i* of infected individuals in both populations to contribute to the force of infection in the other population instead of their own population. An *i* value of 0 corresponds to no interaction, and an *i* value of 0.5 corresponds to complete interaction between the two populations (i.e. is equivalent to one large population). Next, for each of the scenarios above, we additionally consider the impact of varying *R*_0_ between 2 and 16, allowing for improved understanding across a variety of pathogens or viral variants (McMorrow, 2021) (Figure). Finally, we relax the assumption of 95% vaccine efficacy by modelling a range of values from 50% to 90% (Figure S8, S9). Full model equations are shown in SI Appendix A.2. All analyses were conducted in R version 4.0.3.

## 5 Results

### 5.1 Literature review

We reviewed the literature on optimal vaccine allocation across populations that was published prior to the emergence of SARS-CoV-2 (see SI Table A.3.1). Multiple papers (Forster & Gilligan, 2007; Keeling & Shattock, 2012; Klepac et al., 2011; Rowthorn et al., 2009; J. T. Wu et al., 2007) have shown that allocation proportional to population size is rarely optimal. Further, previous studies have highlighted that the timing of vaccine allocation (Matrajt & Longini, 2010; Mylius et al., 2008; Teytelman & Larson, 2013), heterogeneity in population composition, as well as the stochasticity in infection dynamics affect the optimal distribution (Nguyen & Carlson, 2016; Yuan et al., 2015). Duijzer et al. (Duijzer et al., 2018) provide important contributions by showing that the optimal vaccination threshold is often not the herd immunity threshold as further detailed in SI Appendix A.3.2.

### 5.2 Optimal allocation in two populations of equal size

We build upon the existing literature by first examining allocation decisions in the simple scenario of two identical, non-interacting populations with no underlying immune protection to the pathogen (see Figure 1). In the simplest case, with a small number of vaccine doses available, pro-rata allocation performs comparably to highly unequal allocation strategies. As the number of vaccine doses increases, highly unequal strategies gain advantage over pro-rata allocation. This occurs because one population can be vaccinated close to, but lower than, its herd immunity threshold, maximizing the indirect effect of the vaccine doses (Duijzer et al., 2018). When sufficient vaccines are available for both populations to reach that threshold, more unequal strategies use the doses less efficiently, as indicated by the increasing arms of the “W” shapes in Figure 1. Allocating doses to the population that has reached its threshold provides limited benefit in that population and withholds doses from the other. When there are nearly enough doses to reach the thresholds in both populations, the optimal strategy becomes equal allocation between the two populations.

**Figure 1:**
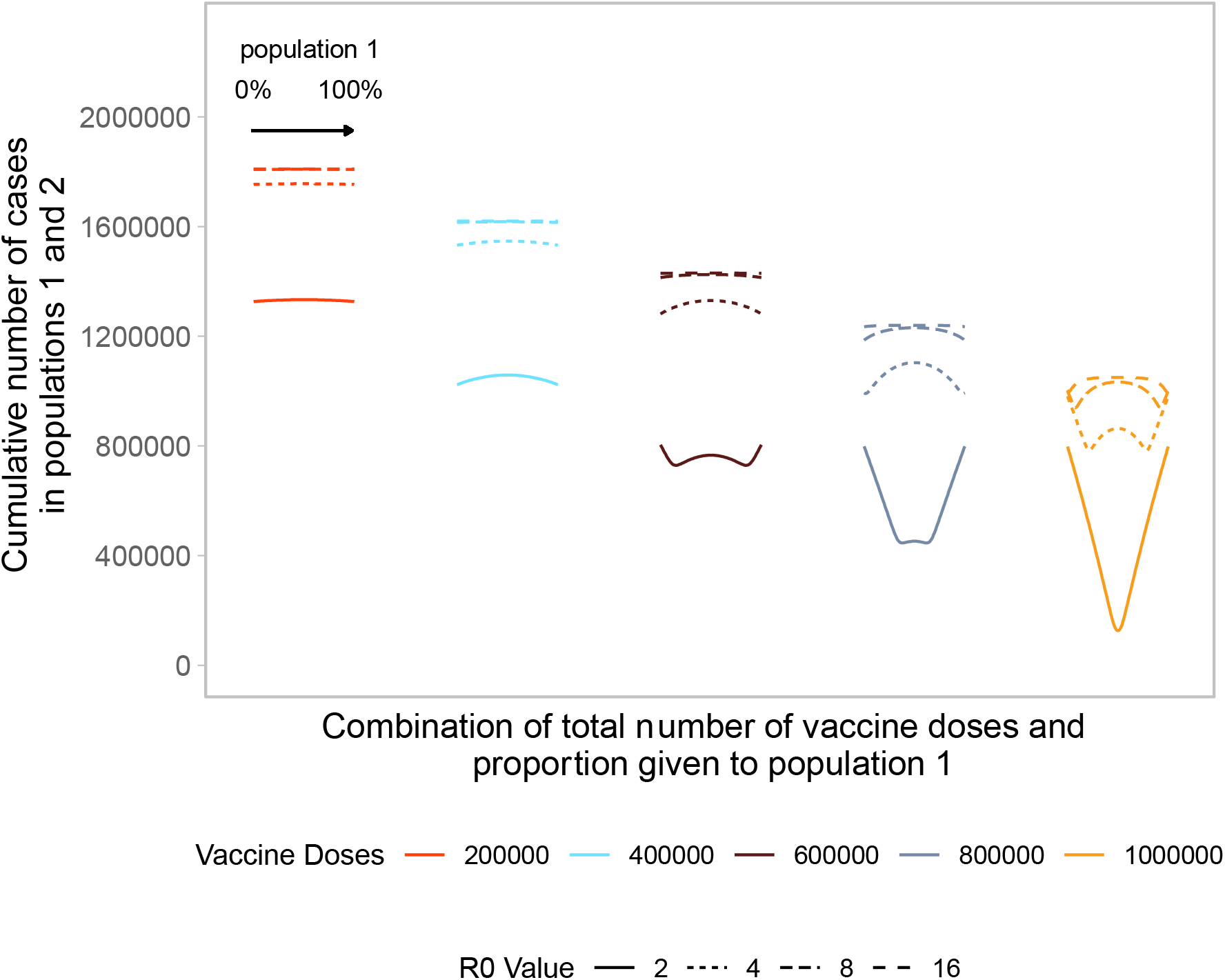
Performance of different allocation strategies of a limited vaccine stockpile across two homogeneous population of equal size with no underlying immunity and prophylactic vaccination. Both populations have one million individuals. Each color represents a different number of total vaccine doses. Each line represents a different basic reproductive number. Each curve shows the cumulative number of cases across both population 1 and 2 for different proportions of doses given each population. Across each curve, from left to right, the proportion of doses to population 1 goes from 0 to 100%. Conversely, for population 2, the proportion of doses goes from 100% to 0%.

As the basic reproductive number increases, we again see that unequal allocations preform optimally, as the number of available doses is less than the number needed to reach the critical herd immunity threshold in both populations. In these scenarios, vaccinating one or the other population until it can reach the critical herd immunity threshold results in the lowest cumulative cases across both populations. At very high basic reproductive numbers (i.e., *R*_0_ = 16), pro-rata allocation performs comparably to highly unequal allocation strategies.

### 5.3 Optimal allocation in two populations of unequal size

Extending the simple case of non-interacting populations of equal size, previous studies have shown how optimal allocation across populations of different sizes is not linear, but varies with the number of doses available in a characteristic, and often counter-intuitive, “switching” pattern (Duijzer et al., 2018; Keeling & Shattock, 2012; Klepac et al., 2011).

As shown in Figure 2 (top), when the number of doses available is very limited, optimal allocation concentrates all vaccine doses to the smallest population, not assigning any to the largest population (regime 1). As the number of doses allocated to the smaller population reaches its threshold, additional doses are gradually allocated to the larger population (regime 2). Strikingly, a drastic switch happens between regimes 2 and 3, and in regime 3 all doses are allocated to the larger population and none to the smaller one. Then, as the largest population itself reaches its threshold, supplementary doses are assigned to the smaller population (regime 4). When the number of vaccines available allows both populations to attain their respective thresholds, vaccines are allocated proportionally to the population sizes (regime 5). Note that here we assume that the total number of doses is fixed at the time of allocation and no additional doses become available over time. We relax this assumption in later scenarios considering continuous rollout.

**Figure 2:**
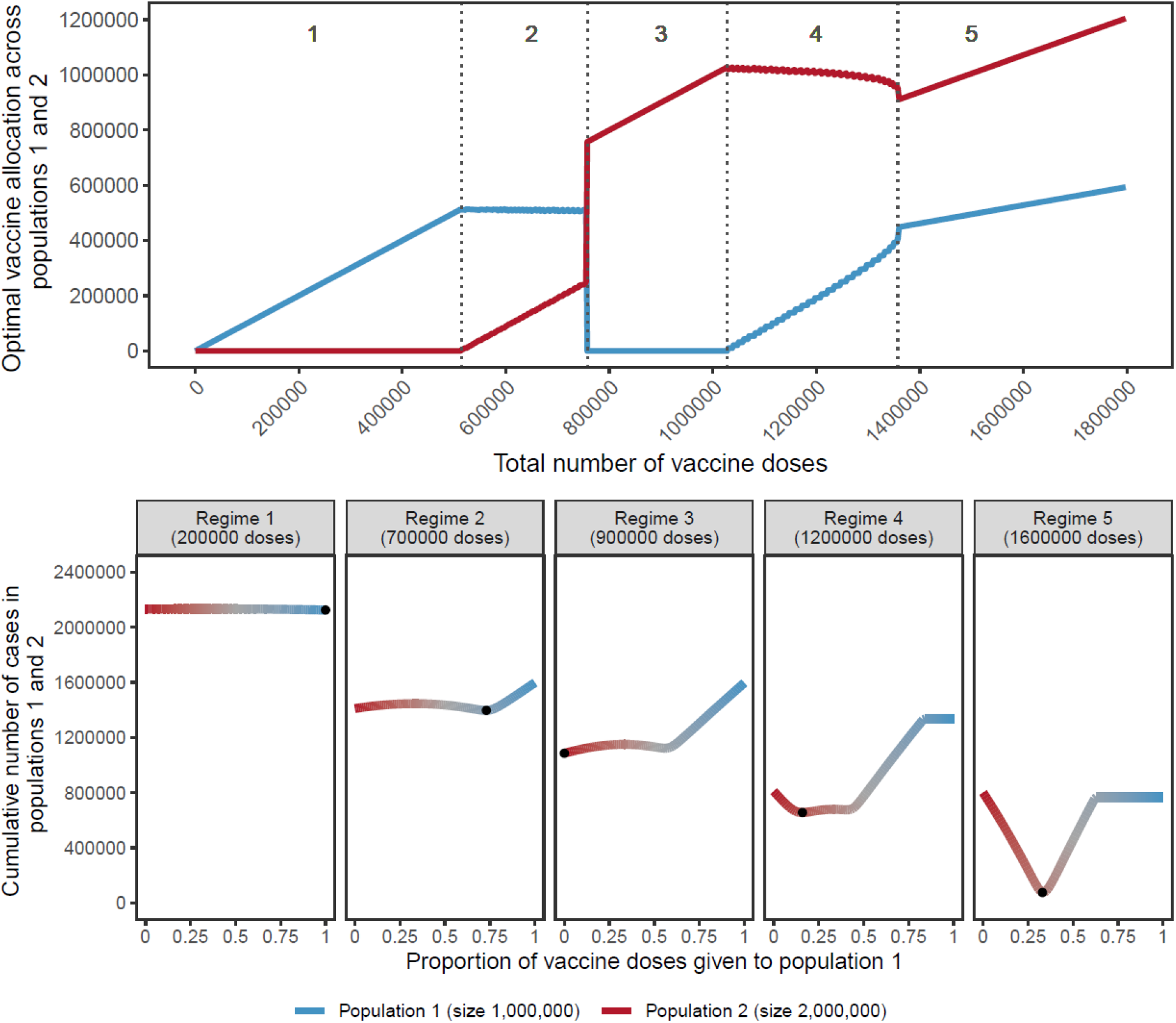
Top: Optimal allocation strategies of a limited vaccine stockpile across two homogeneous populations of unequal size with no underlying immunity, prophylactic vaccination and an R_0_ of 2. Populations 1 (blue) and 2 (red) have one and two million individuals, respectively. Dashed vertical lines were added to highlight regimes (1 to 5) showing different vaccine allocation patterns. Bottom: Performance of allocation strategies for five different numbers of vaccine doses, representative of the regimes shown in the top half of the Figure. Color coding corresponds to vaccine allocation ranging from giving all doses to population 2 (red) to giving all doses to population 1 (blue). The optimal allocation, the minimal value on each plot, is highlighted by a black point.

For most values of vaccine available, the optimal allocation is highly unequal (regime 1, 2, 3, 4), as previously shown (Duijzer et al., 2018; Keeling & Shattock, 2012). This counterintuitive result is caused by the non-linearity of the indirect effect from each additional vaccine dose. Additional doses are allocated to the population where they have the largest benefit. For example, in regime 1 of Figure 2, additional doses bring a larger benefit in the smaller population then they would in the larger population.

Importantly, while prior literature (Keeling & Shattock, 2012) demonstrates that unequal allocations can be optimal, these results show that the benefit of such unequal, optimal allocations over more nearly equal ones is often small. As shown in Figure 2 (bottom), for low numbers of vaccine doses (regimes 1 and 2), although concentrating all doses to the smallest population is optimal, other strategies do not perform much worse. Each regime is characterized by a different allocation profile that gives rise to a different optimum, indicated by black points. In regime 4, the characteristic W shape appears where a fully unequal allocation is sub-optimal, regardless of which population is vaccinated.

### 5.4 Impact of underlying immunity

As vaccines become available to different locations at different points in their local epidemic, populations will have varying degrees of underlying immunity to the virus due to prior infections. Serological surveys estimate that at the end of 2020, around a fifth of the population had already been infected in areas hardest hit during the spring of 2020 (23% in NYC (Rosenberg et al., 2020), 18% in London (Public Health England, 2020) and 11% in Madrid and Paris (Pollán et al., 2020; Salje et al., 2020)). More recent estimates show seroprevalence increased to almost 60% after the Omicron variant became predominant in the United States (Clarke et al., 2022). Select high-risk groups, including health care workers and nursing home residents, have been shown to have an even higher prevalence of SARS-CoV-2 antibodies (Ladhani et al., 2020). To account for underlying immunity, we further simulate optimal allocation decisions with varying levels of underlying immunity in each population to mirror the fact that allocation decisions are made during an ongoing pandemic.

Comparing two populations with varying amounts of underlying immunity, the optimal strategy favors prioritizing the population that is closer to their herd immunity threshold (Figure 3). Figure 3 shows optimal allocation decisions across two homogeneous populations of equal size with no immunity (top left, repeating Figure 1) or increasing degrees of immunity in population 1. With increasing immunity in population 1, the characteristic V- or W-shape becomes more lopsided as fewer doses are required in population 1 to reach the threshold at which doses should be split between populations. Extremely unequal allocation strategies either waste doses or fail to minimize the cumulative number of infections in both populations if given completely to population 1 or 2, respectively. In addition, allocating vaccines to population 1 beyond the amount needed to reach its threshold results in the highest cumulative number of cases because it confers little additional benefit in population 1, and deprives population 2 of vaccines needed to mitigate cases. As before, once the number of doses is large enough to approach or reach the threshold in both populations, optimal strategies move closer to pro-rata allocations.

**Figure 3:**
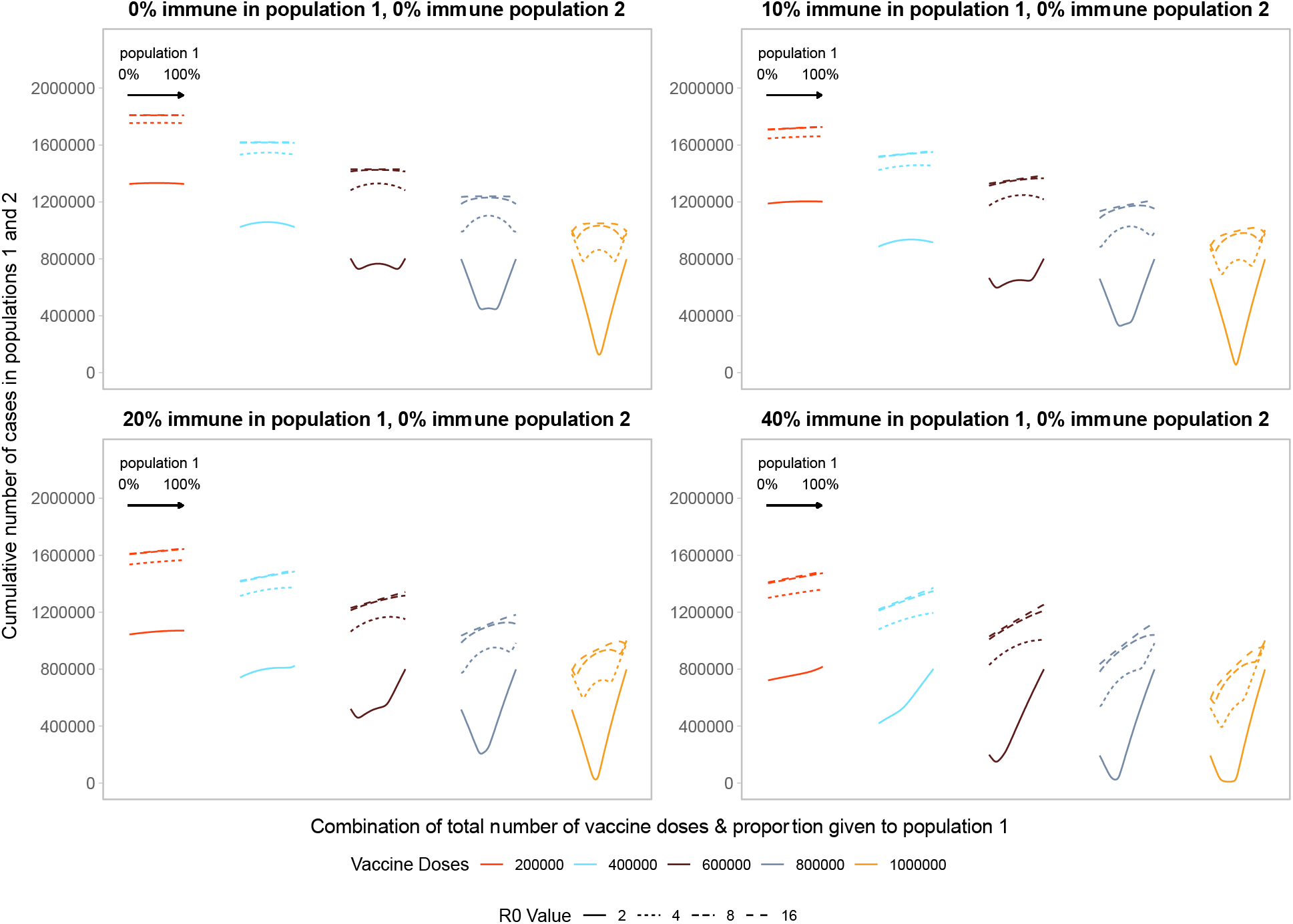
Performance of different allocation strategies of a limited vaccine stockpile across two homogeneous populations of equal size (one million individuals) with different underlying immunity, and prophylactic vaccination. We fix population 2 to have no underlying pathogen immunity and vary underlying immunity in population 1 from 0 to 40%. Each color represents a different number of total vaccine doses. Each line represents a different basic reproductive number. The panel on the top left is equivalent to Figure 1.

As we vary the basic reproductive number, holding vaccine doses fixed, we find the characteristic V and W shapes are shifted to the left. The number of vaccine doses needed to reach the critical herd immunity threshold increases as the basic reproductive number increases. Unequal approaches become more favorable as the level of underlying immunity in population 1 increases, because fewer doses are required for population 1 to reach their herd immunity threshold. Thus, even for very high *R*_0_ values, the optimal strategy, minimizing the cumulative number of cases across both populations, prioritizes allocating doses to the population that is closest to reaching its critical herd immunity threshold.

### 5.5 Impact of delayed vaccine roll-out in a homogeneous population

Next, we examine the impact of vaccine roll-out over the course of the epidemic. We find both the timing and speed of roll-out play an important role in minimizing the final size of the epidemic. As shown in Figure 4, the cumulative number of cases across both populations is minimized when vaccine roll-out occurs as soon as possible after the start of the epidemic. Further, the final size is minimized when roll-out speed is increased, vaccinating a larger proportion of the population each day.

**Figure 4:**
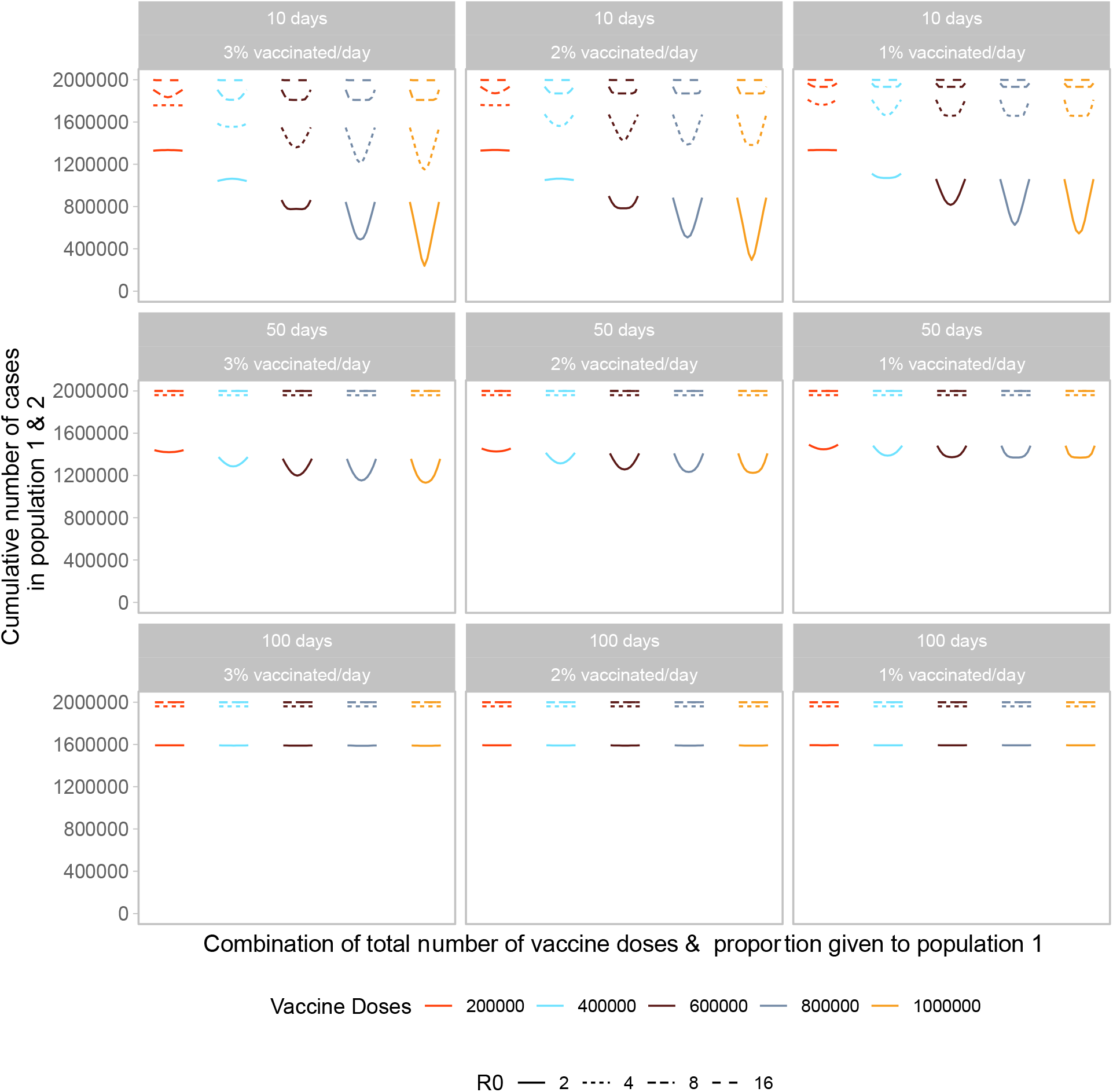
Performance of different allocation strategies of a limited vaccine stockpile across two homogeneous populations of equal size (one million individuals) with no underlying immunity, with vaccines rolled out at different speeds and different times after the start of the epidemic. Each color represents a different number of total vaccine doses. Each line represents a different basic reproductive number. We vary both the timing and speed of roll-out between 10, 50 or 100 days after the start of the epidemic with 1, 2, or 3% of the population vaccinated per day. Each column represents a given roll-out speed while each row represents a different timing.

For the early and efficient roll-out (beginning 10 days after the start of the epidemic, at a rate of 2 or 3% of the population/day), the vaccination performance profile across possible allocations looks similar to that of the prophylactic vaccine deployment strategy shown in Figure 1. However, for a slower or more delayed roll-out we see highly unequal approaches perform poorly across almost all doses and more equal approaches result in the smallest final size. This is because a larger fraction of the population is naturally infected, minimizing the gains from concentrating vaccine doses in one population.

As we incorporate differences in transmissibility, we find timing and speed to be of greater importance. Even with a vaccine roll-out 50 days after the start of the epidemic, there are no differences in final size across all allocation strategies, within a given *R*_0_ level, as the epidemic has ended in the population before vaccines are introduced. For higher reproductive numbers, faster, earlier roll-outs are needed for vaccination to have an impact on the total number of infections.

### 5.6 Impact of heterogeneous population structure

Looking within a population, many studies have shown optimal strategies favor prioritizing older individuals (e.g., those aged 60 or over) or those with certain comorbidities when the goal is minimizing mortality. If the goal instead is minimizing final size, targeting adults 20-49 with an effective transmission-blocking vaccine minimizes cumulative incidence (Bubar et al., 2021; Matrajt et al., 2021). Here we model the impact of heterogeneous population structure to examine the impact of strategies across populations. These simulations consider populations with heterogeneous transmission or with heterogeneous risk of death.

Targeting high transmission or high mortality groups first within a population shifts the optimal allocation across the two populations towards pro-rata allocation (Figure 5, S3). In Figure 5 we first model the impact of prophylactic vaccination in a heterogeneous population structure with 25% of each population at either high risk of transmission (top) or death (bottom). In the high-transmission scenario, the behavior looks similar to that in Figure 1 for a low number of doses, representing the trade-off between vaccinating the high-transmitters in both populations. Once there are enough doses available to vaccinate enough high-transmitters to reduce transmission dramatically, the optimal strategy favors more pro-rata allocations across the two populations as high-transmitters are driving the bulk of transmission. This shift to more equal allocations occurs at a lower number of vaccine doses compared to Figure 1. In the high-mortality scenario, we see the optimal allocation rapidly shift to pro-rata strategies, starting at a very low number of vaccine doses. Interestingly, the sequence of profiles from Figure 1 is repeated twice. First, for a low number of vaccine doses there is a trade-off between vaccinating the high-mortality individuals in both populations. Then for higher vaccine counts the trade-off is repeated, this time between all individuals of both populations. While this trade-off exists, pro-rata allocation is heavily favored across almost all levels of available vaccine doses.

**Figure 5:**
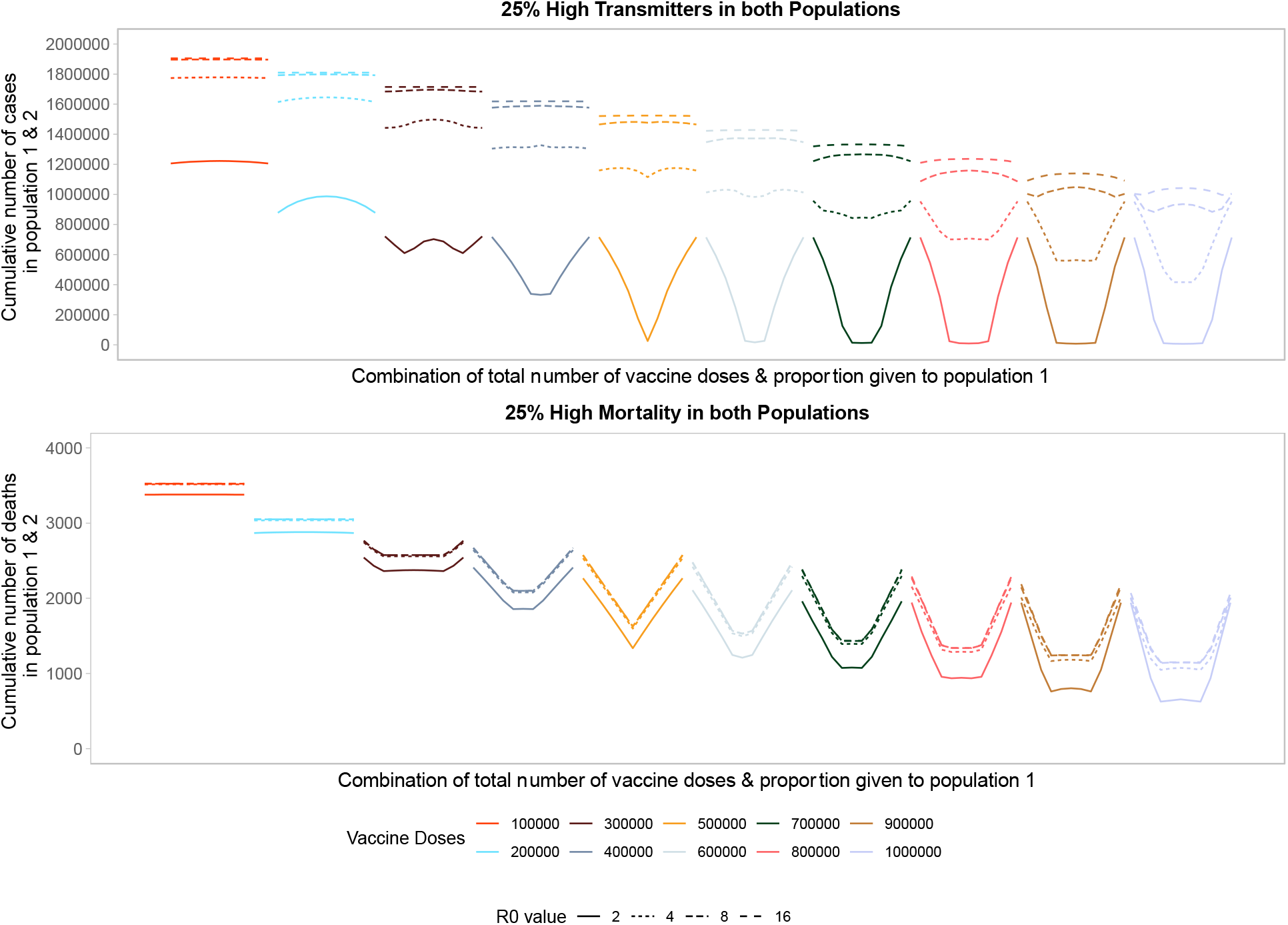
Performance of different allocation strategies of a limited vaccine stockpile across two heterogeneous populations of equal size (one million individuals) with no underlying immunity and prophylactic vaccination. Each color represents a different number of total vaccine doses. Each line represents a different basic reproductive number. In both the high transmission scenario (top) and high mortality scenario (bottom), 25% of both populations are high risk.

Looking across different levels of *R*_0_, we find similar trends. Vaccinating higher transmission or mortality groups first results in more equal allocation strategies across populations. For higher *R*_0_ values (i.e., 8 or 16) pro-rata allocation performs comparably to highly unequal strategies. Increasing the proportion of high-risk individuals to 50% of the population (Figure S3) we find similar trends for *R*_0_ values of 2 and 4. For higher *R*_0_ values, unequal approaches perform optimally as a larger fraction of the population is driving transmission, so effectively targeting this group in either population minimizes the cumulative number or deaths or cases across the two populations.

Next, we considered the impact of continuous roll-out for both the high transmission and high mortality scenarios. We find that across both high-risk scenarios and all vaccine roll-out times and speeds, unequal allocation is highly sub-optimal (Figure S4-S7). Similarly to Figure 4, we vary the start date of vaccination roll-out (1, 10, 30, 50, or 100 days), the daily vaccination rate (1, 2 or 3% per day), and the proportion of the population at high risk (25 or 50%). We find that both the speed and timing of vaccine roll-out are important factors in minimizing the cumulative number of cases or deaths across the two populations and see the greatest reduction in cumulative deaths and final size with the earliest and fastest roll-out. Specifically, for vaccine stockpiles larger than 500,000 doses, the achievable impact of vaccination is more dependent on the timing (solid vs. dashed curves) and speed (different panels) of vaccine roll-out rather than on the total number of doses available.

### 5.7 Sensitivity Analyses

We assessed the robustness of our results by varying the characteristics of the vaccine and connection between populations to be more representative of the current pandemic. As expected, the leaky and all-or-nothing vaccine have the same critical vaccination threshold, though the cumulative number of cases in the leaky vaccine scenario is equal to or larger than the all-or - nothing scenario (Magpantay et al., 2014) (Figure S1).

In the previous situations we have only considered the scenario of non-interacting populations. As we relax this strict assumption, we find that as the amount of interaction between the two populations increases, equal strategies are most favorable (Figure S2). When the force of infection in each population depends on epidemic dynamics in both populations, accounting for interaction drastically changes the optimal allocation profiles and favors equal allocation between populations, as seen in previous work (Duijzer et al., 2018; Keeling & Shattock, 2012). Even for low values of the interaction parameter *i*, equal allocation rapidly becomes optimal. Indeed, for *i* values higher than 0.01 — which corresponds to one out of every hundred infected individuals contributing to infection in the other population — equal allocation between the two population always performs best. As *i* further increases, unequal strategies progressively approach the optimal (equal) allocation as indicated by the flattening of the curves. For *i* equal to 0.5, when the two populations concretely behave like one large population, all allocation strategies perform almost identically. Compared to the non-interacting case, allowing for interaction between the two populations leads to a higher cumulative number of infections for all possible vaccine allocation strategies, and the “W”-shaped allocation curve no longer appears.

As we increase the basic reproductive number, we find that the optimal strategy quickly favors more equal allocation decisions. In addition, interaction between the two populations becomes less important for very high values of R_0_, as the allocation profiles look similar across all interaction parameters for R_0_ values of 8 and 16.

As vaccine efficacy decreases from 90% to 50%, the cumulative number of cases across both populations increases, the critical herd immunity threshold increases and equal-allocation strategies become less favorable, with unequal allocations being optimal in some cases for the lowest efficacy values. Even for these situations, however, the advantage of unequal allocations are modest (Figure S8). Next, as we increase immunity levels across both populations, the total number of cases is reduced, and the critical herd immunity threshold is lowered. Even with low vaccine efficacy values, in scenarios with high underlying immunity, the critical herd immunity threshold can be reached, and equal allocations are favored (Figure S9).

## 6 Discussion

In emerging pandemics, countries must make challenging vaccine allocation decisions due to resource constraints. Previous studies (Keeling & Shattock, 2012) have shown simple scenarios favor unequal allocation. We recreated those findings, and further extend vaccine allocation theory, and apply it to scenarios similar to the 2019 SARS-CoV-2 and 2009 H1N1 influenza pandemics. We focus on these emerging pathogens as those pandemics are the ones for which we have the greatest amount of data, understanding, and for which vaccines were deployed while the pandemic was ongoing.

In the simple case of two non-interacting populations of identical size we show that for very high quantities of vaccine, relative to population size, equal allocation strategies are optimal. For very few doses, all strategies provide comparable results. This supports the European Commission’s decision to allocate vaccine doses proportional to population size among the 27 European Nations (European Commission, 2020). In this simplest model, until there is enough vaccine for both populations to approach their critical herd immunity threshold, optimal strategies favor a highly unequal approach, allocating doses to either population 1 or 2 until the population has reached its threshold. If the populations vary in size, allocation decisions vary, and as the number of vaccines increases, focus switches from the smaller population to the larger one, as supported by Keeling and Shattock (Keeling & Shattock, 2012).

We consider more realistic scenarios that better mirror the diversity and complexities of emerging pandemics including underlying immunity, population interaction, continuous vaccine roll-out, heterogeneous population structure, and differences in underlying disease transmissibility (either because of biological or social factors). While many of these parameters are either unknown or changing throughout the course of the epidemic, we find that, across a range of scenarios, optimal allocation decisions often favor equal allocation across populations. Since these strategies are often optimal or nearly optimal across a range of parameters, while unequal allocations are only generally optimal for narrow parameter ranges, more pro-rata strategies might be the best option under uncertainty in an ongoing epidemic. Moreover, during an emerging pandemic, it may be unclear whether the newly developed vaccines confer protection against transmission, thus limiting the potential benefit from unequal vaccine allocation strategies that rely on maximizing the benefit from indirect protection in one population. It may thus be preferable to focus on equal allocation strategies as those rely more on the direct protection against disease. Parameter values from COVID-19 and pandemic influenza illustrate this phenomenon; however, these results contribute more generally to the existing vaccine allocation theory for any epidemic emerging in multiple populations when key epidemic variables remain unknown.

For scenarios considering heterogeneous population risk, we find that first targeting high risk individuals, either high-transmitters or those at higher risk of death after infection, results in more equal allocations between populations being optimal. Targeting high-risk individuals first, then shifting priority to lower-risk individuals is supported by previous modeling work, looking at SARS-CoV-2 vaccine allocations within a single population (Bubar et al., 2021; Chen et al., 2020; Hogan et al., 2020; Matrajt et al., 2021; Moore et al., 2021), and is in concordance with the ongoing COVAX strategy, targeting early doses to high-risk individuals, and the USA’s implement which vaccinated health care workers and elderly individuals first (Gayle et al., 2020; World Health Organization, 2020). This also supports previous guidelines for 2009 H1N1 influenza where vaccines were targeted at high risk groups first, before shifting to the general population (Centers for Disease Control and Prevention, 2009).

Our modeling analyses are subject to many simplifying assumptions on population dynamics and vaccine characteristics that may not be applicable to the current pandemic. We consider a vaccine that prevents both disease and infection, thus providing indirect protection to a fraction of the population. While some vaccines are able to reduce infectiousness, in future pandemics this effect will still need to be precisely assessed. We do not model vaccine refusal and assume that all individuals given doses accept them. Recent studies (Dror et al., 2020) show vaccine hesitancy as a threat to successful pandemic response. Next, we do not consider delays between doses, but model the epidemic from a final dose which confers 95% efficacy. Due to vaccine shortages, the delay between the first and second dose could impact our findings as individuals may be able to get infected in the interim. Further, we only model one available vaccine. The current SARS-CoV-2 pandemic illustrates how the vaccine landscape can be complex, as multiple vaccines are available, with many more rapidly undergoing testing. Considerations for optimal allocation in this context are more complicated, especially if the vaccines have different immunogenicity profiles, and the quantity of doses and timing of roll-out varies across candidates. Finally, we do not consider the impact of non-pharmaceutical interventions (NPIs) in conjunction with vaccination.

Furthermore, we only model allocation strategies within two symmetric populations. It is likely that policy makers will face allocation decisions across multiple countries, or across multiple regions within a country. While our analyses do not extend to more than two locations, general principles should remain the same, as illustrated elsewhere for three populations (Keeling & Shattock, 2012),(Duijzer et al., 2018).

Future modeling work on vaccination strategies during emerging pandemics is needed, for example considering scenarios where multiple vaccine candidates are rolled out simultaneously. These studies should also consider the effects of vaccines on reducing hospitalizations and preserving hospital capacity, which may have indirect benefits for mortality rates for COVID-19 and other diseases beyond the direct prevention of infection in high-risk populations. In addition, other work should also consider populations with varying epidemic dynamics, and distribution capacity. Indeed, it has been argued that populations at higher immediate risk of disease spread and populations where vaccine roll-out is most efficient should be prioritized for vaccine allocation (Emanuel & Persad, 2021).

With vaccine supplies usually severely constrained, in the future rapid allocation decisions will need to be made while the pandemic is ongoing. Due to the global impact and magnitude of some pandemics such as the current 2019 SARS-CoV-2 pandemic, further political and economic constraints will likely play a large role in allocation decisions. Mathematical modelling can provide insight into optimal allocation strategies that maximize the benefit from each dose. Conclusions from such models should be balanced with ethical considerations on the fairness of allocation that also minimize disparities in access. We show key principles that should be considered in the design of realistic and implementable allocation strategies.

## Data Availability

All relevant data and code will be available on GitHub.

## 7 Acknowledgments

We thank Dr. Kendall Hoyt for helpful discussions.

## References

Bubar, K. M., Reinholt, K., Kissler, S. M., Lipsitch, M., Cobey, S., Grad, Y. H., & Larremore, D. B. (2021). Model-informed COVID-19 vaccine prioritization strategies by age and serostatus. Science, 371(6532), 916–921. https://doi.org/10.1126/science.abe6959

Buckner, J. H., Chowell, G., & Springborn, M. R. (2021). Dynamic prioritization of COVID-19 vaccines when social distancing is limited for essential workers. Proceedings of the National Academy of Sciences, 118(16). https://doi.org/10.1073/pnas.2025786118

Centers for Disease Control and Prevention. (2009). 2009 H1N1 Vaccine Recommendations. https://www.cdc.gov/h1n1flu/vaccination/acip.htm

Chen, X., Li, M., Simchi-Levi, D., & Zhao, T. (2020). Allocation of COVID-19 Vaccines Under Limited Supply. MedRxiv, 2020.08.23.20179820. https://doi.org/10.1101/2020.08.23.20179820

Chowell, G., Viboud, C., Wang, X., Bertozzi, S. M., & Miller, M. A. (2009). Adaptive Vaccination Strategies to Mitigate Pandemic Influenza: Mexico as a Case Study. PLoS ONE, 4(12), e8164. https://doi.org/10.1371/journal.pone.0008164

Clarke, K. E. N., Jones, J. M., Deng, Y., Nycz, E., Lee, A., Iachan, R., Gundlapalli, A. V., Hall, A. J., & MacNeil, A. (2022). Seroprevalence of Infection-Induced SARS-CoV-2 Antibodies — United States, September 2021–February 2022. MMWR. Morbidity and Mortality Weekly Report, 71(17), 606–608. https://doi.org/10.15585/mmwr.mm7117e3

Dawood, F. S., Iuliano, A. D., Reed, C., Meltzer, M. I., Shay, D. K., Cheng, P.-Y., Bandaranayake, D., Breiman, R. F., Brooks, W. A., Buchy, P., Feikin, D. R., Fowler, K. B., Gordon, A., Hien, N. T., Horby, P., Huang, Q. S., Katz, M. A., Krishnan, A., Lal, R., … Widdowson, M.-A. (2012). Estimated global mortality associated with the first 12 months of 2009 pandemic influenza A H1N1 virus circulation: a modelling study. The Lancet Infectious Diseases, 12(9), 687–695. https://doi.org/10.1016/S1473-3099(12)70121-4

Dong, E., Du, H., & Gardner, L. (2020). An interactive web-based dashboard to track COVID-19 in real time. The Lancet Infectious Diseases, 20(5), 533–534. https://doi.org/10.1016/S1473-3099(20)30120-1

Doria-Rose, N., Suthar, M. S., Makowski, M., O’Connell, S., McDermott, A. B., Flach, B., Ledgerwood, J. E., Mascola, J. R., Graham, B. S., Lin, B. C., O’Dell, S., Schmidt, S. D., Widge, A. T., Edara, V.-V., Anderson, E. J., Lai, L., Floyd, K., Rouphael, N. G., Zarnitsyna, V., … Kunwar, P. (2021). Antibody Persistence through 6 Months after the Second Dose of mRNA-1273 Vaccine for Covid-19. New England Journal of Medicine, 384(23), 2259–2261. https://doi.org/10.1056/NEJMc2103916

Dror, A. A., Eisenbach, N., Taiber, S., Morozov, N. G., Mizrachi, M., Zigron, A., Srouji, S., & Sela, E. (2020). Vaccine hesitancy: the next challenge in the fight against COVID-19. European Journal of Epidemiology, 35(8), 775–779. https://doi.org/10.1007/s10654-020-00671-y

Duijzer, L. E., van Jaarsveld, W. L., Wallinga, J., & Dekker, R. (2018). Dose-Optimal Vaccine Allocation over Multiple Populations. Production and Operations Management, 27(1), 143–159. https://doi.org/10.1111/poms.12788

Emanuel, E. J., & Persad, G. (2021, May 24). This Is the Wrong Way to Distribute Badly Needed Vaccinese. New York Times. https://www.nytimes.com/2021/05/24/opinion/vaccine-covid-distribution.html

European Commission. (2020). Communication from the Commission EU Strategy for COVID-19 vaccines. https://ec.europa.eu/info/sites/default/files/communication-eu-strategy-vaccines-covid19_en.pdf

Fidler, D. P. (2010). Negotiating Equitable Access to Influenza Vaccines: Global Health Diplomacy and the Controversies Surrounding Avian Influenza H5N1 and Pandemic Influenza H1N1. PLoS Medicine, 7(5), e1000247. https://doi.org/10.1371/journal.pmed.1000247

Fineberg, H. V. (2014). Pandemic Preparedness and Response — Lessons from the H1N1 Influenza of 2009. New England Journal of Medicine, 370(14), 1335–1342. https://doi.org/10.1056/NEJMra1208802

Forster, G. A., & Gilligan, C. A. (2007). Optimizing the control of disease infestations at the landscape scale. Proceedings of the National Academy of Sciences, 104(12), 4984–4989. https://doi.org/10.1073/pnas.0607900104

Gayle, H., Foege, W., Brown, L., & Kahn, B. (Eds.). (2020). Framework for Equitable Allocation of COVID-19 Vaccine. National Academies Press. https://doi.org/10.17226/25917

Goldstein, E., Lipsitch, M., & Cevik, M. (2021). On the Effect of Age on the Transmission of SARS-CoV-2 in Households, Schools, and the Community. The Journal of Infectious Diseases, 223(3), 362–369. https://doi.org/10.1093/infdis/jiaa691

Hogan, A. B., Winskill, P., Watson, O. J., Walker, P. G., Whittaker, C., Baguelin, M., Haw, D., Lochen, A., Gaythorpe, K. A., Team, I. C.-19 R., Muhib, F., Smith, P., Hauck, K., Ferguson, N. M., & Ghani, A. C. (2020). Modelling the allocation and impact of a COVID-19 vaccine. Imperial College London. https://doi.org/10.25561/82822

Islam, M. R., Oraby, T., McCombs, A., Chowdhury, M. M., Al-Mamun, M., Tyshenko, M. G., & Kadelka, C. (2021). Evaluation of the United States COVID-19 vaccine allocation strategy. PLOS ONE, 16(11), e0259700. https://doi.org/10.1371/journal.pone.0259700

Kaiser Family Foundation. (2009). Nine Countries Pledge H1N1 Vaccine Donations To Developing Countries. https://www.kff.org/news-summary/nine-countries-pledge-h1n1-vaccine-donations-to-developing-countries/

Keeling, M. J., & Shattock, A. (2012). Optimal but unequitable prophylactic distribution of vaccine. Epidemics, 4(2), 78–85. https://doi.org/10.1016/j.epidem.2012.03.001

Kissler, S. M., Viboud, C., Grenfell, B. T., & Gog, J. R. (2020). Symbolic transfer entropy reveals the age structure of pandemic influenza transmission from high-volume influenza-like illness data. Journal of The Royal Society Interface, 17(164), 20190628. https://doi.org/10.1098/rsif.2019.0628

Klepac, P., Laxminarayan, R., & Grenfell, B. T. (2011). Synthesizing epidemiological and economic optima for control of immunizing infections. Proceedings of the National Academy of Sciences, 108(34), 14366–14370. https://doi.org/10.1073/pnas.1101694108

Ladhani, S. N., Jeffery-Smith, A., Patel, M., Janarthanan, R., Fok, J., Crawley-Boevey, E., Vusirikala, A., Fernandez Ruiz De Olano, E., Perez, M. S., Tang, S., Dun-Campbell, K., Evans, E. W.-, Bell, A., Patel, B., Amin-Chowdhury, Z., Aiano, F., Paranthaman, K., Ma, T., Saavedra-Campos, M., … Zambon, M. (2020). High prevalence of SARS-CoV-2 antibodies in care homes affected by COVID-19: Prospective cohort study, England. EClinicalMedicine, 28, 100597. https://doi.org/10.1016/j.eclinm.2020.100597

Lee, B. Y., Brown, S. T., Korch, G. W., Cooley, P. C., Zimmerman, R. K., Wheaton, W. D., Zimmer, S. M., Grefenstette, J. J., Bailey, R. R., Assi, T.-M., & Burke, D. S. (2010). A computer simulation of vaccine prioritization, allocation, and rationing during the 2009 H1N1 influenza pandemic. Vaccine, 28(31), 4875–4879. https://doi.org/10.1016/j.vaccine.2010.05.002

Magpantay, F. M. G., Riolo, M. A., de Cellès, M. D., King, A. A., & Rohani, P. (2014). Epidemiological Consequences of Imperfect Vaccines for Immunizing Infections. SIAM Journal on Applied Mathematics, 74(6), 1810–1830. https://doi.org/10.1137/140956695

Mathieu, E., Ritchie, H., Ortiz-Ospina, E., Roser, M., Hasell, J., Appel, C., Giattino, C., & Rodés-Guirao, L. (2021). A global database of COVID-19 vaccinations. Nature Human Behaviour, 5(7), 947–953. https://doi.org/10.1038/s41562-021-01122-8

Matrajt, L., Eaton, J., Leung, T., & Brown, E. R. (2021). Vaccine optimization for COVID-19: Who to vaccinate first? Science Advances, 7(6). https://doi.org/10.1126/sciadv.abf1374

Matrajt, L., & Longini, I. M. (2010). Optimizing Vaccine Allocation at Different Points in Time during an Epidemic. PLoS ONE, 5(11), e13767. https://doi.org/10.1371/journal.pone.0013767

McMorrow, M. (2021). Improving communications around vaccine breakthrough and vaccine effectiveness. https://context-cdn.washingtonpost.com/notes/prod/default/documents/8a726408-07bd-46bd-a945-3af0ae2f3c37/note/57c98604-3b54-44f0-8b44-b148d8f75165

Monod, M., Blenkinsop, A., Xi, X., Hebert, D., Bershan, S., Tietze, S., Baguelin, M., Bradley, V. C., Chen, Y., Coupland, H., Filippi, S., Ish-Horowicz, J., McManus, M., Mellan, T., Gandy, A., Hutchinson, M., Unwin, H. J. T., van Elsland, S. L., Vollmer, M. A. C., … Ratmann, O. (2021). Age groups that sustain resurging COVID-19 epidemics in the United States. Science, 371(6536). https://doi.org/10.1126/science.abe8372

Moore, S., Hill, E. M., Dyson, L., Tildesley, M. J., & Keeling, M. J. (2021). Modelling optimal vaccination strategy for SARS-CoV-2 in the UK. PLOS Computational Biology, 17(5), e1008849. https://doi.org/10.1371/journal.pcbi.1008849

Mylius, S. D., Hagenaars, T. J., Lugnér, A. K., & Wallinga, J. (2008). Optimal allocation of pandemic influenza vaccine depends on age, risk and timing. Vaccine, 26(29–30), 3742–3749. https://doi.org/10.1016/j.vaccine.2008.04.043

Nguyen, C., & Carlson, J. M. (2016). Optimizing Real-Time Vaccine Allocation in a Stochastic SIR Model. PLOS ONE, 11(4), e0152950. https://doi.org/10.1371/journal.pone.0152950

Pollán, M., Pérez-Gómez, B., Pastor-Barriuso, R., Oteo, J., Hernán, M. A., Pérez-Olmeda, M., Sanmartín, J. L., Fernández-García, A., Cruz, I., Fernández de Larrea, N., Molina, M., Rodríguez-Cabrera, F., Martín, M., Merino-Amador, P., León Paniagua, J., Muñoz-Montalvo, J. F., Blanco, F., Yotti, R., Blanco, F., … Vázquez de la Villa, A. (2020). Prevalence of SARS-CoV-2 in Spain (ENE-COVID): a nationwide, population-based seroepidemiological study. The Lancet, 396(10250), 535–544. https://doi.org/10.1016/S0140-6736(20)31483-5

Presanis, A. M., De Angelis, D., Team3¶, T. N. Y. C. S. F. I., Hagy, A., Reed, C., Riley, S., Cooper, B. S., Finelli, L., Biedrzycki, P., & Lipsitch, M. (2009). The Severity of Pandemic H1N1 Influenza in the United States, from April to July 2009: A Bayesian Analysis. PLOS Medicine, 6(12), e1000207. https://doi.org/10.1371/journal.pmed.1000207

Public Health England. (2020). Sero-surveillance of COVID-19. https://assets.publishing.service.gov.uk/government/uploads/system/uploads/attachment_data/file/888254/COVID19_Epidemiological_Summary_w22_Final.pdf

Rosenberg, E. S., Tesoriero, J. M., Rosenthal, E. M., Chung, R., Barranco, M. A., Styer, L. M., Parker, M. M., John Leung, S.-Y., Morne, J. E., Greene, D., Holtgrave, D. R., Hoefer, D., Kumar, J., Udo, T., Hutton, B., & Zucker, H. A. (2020). Cumulative incidence and diagnosis of SARS-CoV-2 infection in New York. Annals of Epidemiology, 48, 23-29.e4. https://doi.org/10.1016/j.annepidem.2020.06.004

Rowthorn, R. E., Laxminarayan, R., & Gilligan, C. A. (2009). Optimal control of epidemics in metapopulations. Journal of The Royal Society Interface, 6(41), 1135–1144. https://doi.org/10.1098/rsif.2008.0402

Rydland, H. T., Friedman, J., Stringhini, S., Link, B. G., & Eikemo, T. A. (2022). The radically unequal distribution of Covid-19 vaccinations: a predictable yet avoidable symptom of the fundamental causes of inequality. Humanities and Social Sciences Communications, 9(1), 61. https://doi.org/10.1057/s41599-022-01073-z

Salje, H., Tran Kiem, C., Lefrancq, N., Courtejoie, N., Bosetti, P., Paireau, J., Andronico, A., Hozé, N., Richet, J., Dubost, C.-L., Le Strat, Y., Lessler, J., Levy-Bruhl, D., Fontanet, A., Opatowski, L., Boelle, P.-Y., & Cauchemez, S. (2020). Estimating the burden of SARS-CoV-2 in France. Science, 369(6500), 208–211. https://doi.org/10.1126/science.abc3517

Simonsen, L., Spreeuwenberg, P., Lustig, R., Taylor, R. J., Fleming, D. M., Kroneman, M., Van Kerkhove, M. D., Mounts, A. W., & Paget, W. J. (2013). Global Mortality Estimates for the 2009 Influenza Pandemic from the GLaMOR Project: A Modeling Study. PLoS Medicine, 10(11), e1001558. https://doi.org/10.1371/journal.pmed.1001558

Teytelman, A., & Larson, R. C. (2013). Multiregional Dynamic Vaccine Allocation During an Influenza Epidemic. Service Science, 5(3), 197–215. https://doi.org/10.1287/serv.2013.0046

Tuite, A. R., Fisman, D. N., Kwong, J. C., & Greer, A. L. (2010). Optimal Pandemic Influenza Vaccine Allocation Strategies for the Canadian Population. PLoS ONE, 5(5), e10520. https://doi.org/10.1371/journal.pone.0010520

Watson, O. J., Barnsley, G., Toor, J., Hogan, A. B., Winskill, P., & Ghani, A. C. (2022). Global impact of the first year of COVID-19 vaccination: a mathematical modelling study. The Lancet Infectious Diseases. https://doi.org/10.1016/S1473-3099(22)00320-6

Williamson, E. J., Walker, A. J., Bhaskaran, K., Bacon, S., Bates, C., Morton, C. E., Curtis, H. J., Mehrkar, A., Evans, D., Inglesby, P., Cockburn, J., McDonald, H. I., MacKenna, B., Tomlinson, L., Douglas, I. J., Rentsch, C. T., Mathur, R., Wong, A. Y. S., Grieve, R., … Goldacre, B. (2020). Factors associated with COVID-19-related death using OpenSAFELY. Nature, 584(7821), 430–436. https://doi.org/10.1038/s41586-020-2521-4

World Health Organization. (2011). Pandemic influenza preparedness Framework for the sharing of influenza viruses and access to vaccines and other benefits. https://apps.who.int/gb/pip/pdf_files/pandemic-influenza-preparedness-en.pdf

World Health Organization. (2020). WHO Concept for fair access and equitable allocation of COVID19 health products. https://www.who.int/docs/default-source/coronaviruse/who-covid19-vaccine-allocation-final-working-version-9sept.pdf

Wu, C., Chen, X., Cai, Y., Xia, J., Zhou, X., Xu, S., Huang, H., Zhang, L., Zhou, X., Du, C., Zhang, Y., Song, J., Wang, S., Chao, Y., Yang, Z., Xu, J., Zhou, X., Chen, D., Xiong, W., … Song, Y. (2020). Risk Factors Associated With Acute Respiratory Distress Syndrome and Death in Patients With Coronavirus Disease 2019 Pneumonia in Wuhan, China. JAMA Internal Medicine, 180(7), 934. https://doi.org/10.1001/jamainternmed.2020.0994

Wu, J. T., Riley, S., & Leung, G. M. (2007). Spatial considerations for the allocation of pre-pandemic influenza vaccination in the United States. Proceedings of the Royal Society B: Biological Sciences, 274(1627), 2811–2817. https://doi.org/10.1098/rspb.2007.0893

Yuan, E. C., Alderson, D. L., Stromberg, S., & Carlson, J. M. (2015). Optimal Vaccination in a Stochastic Epidemic Model of Two Non-Interacting Populations. PLOS ONE, 10(2), e0115826. https://doi.org/10.1371/journal.pone.0115826

Zhou, F., Yu, T., Du, R., Fan, G., Liu, Y., Liu, Z., Xiang, J., Wang, Y., Song, B., Gu, X., Guan, L., Wei, Y., Li, H., Wu, X., Xu, J., Tu, S., Zhang, Y., Chen, H., & Cao, B. (2020). Clinical course and risk factors for mortality of adult inpatients with COVID-19 in Wuhan, China: a retrospective cohort study. The Lancet, 395(10229), 1054–1062. https://doi.org/10.1016/S0140-6736(20)30566-3

