## Supporting Information for "Comparative performance of between-population vaccine allocation strategies with applications for emerging pandemics"

Keya Joshi[1,*,a], Eva Rumpler[1,*,a], Lee Kennedy-Shaffer[a,b], Rafia Bosan[a] and Marc Lipsitch[a]

[a] Center for Communicable Disease Dynamics, Department of Epidemiology, Harvard TH Chan School of Public Health, 02115 Boston, Massachusetts

[b] Department of Mathematics & Statistics, Vassar College, 12604 Poughkeepsie, New York

[1]Contributed equally to this work.
[*]Corresponding authors.

### A Supporting Information

#### A.1 Supplementary Figures

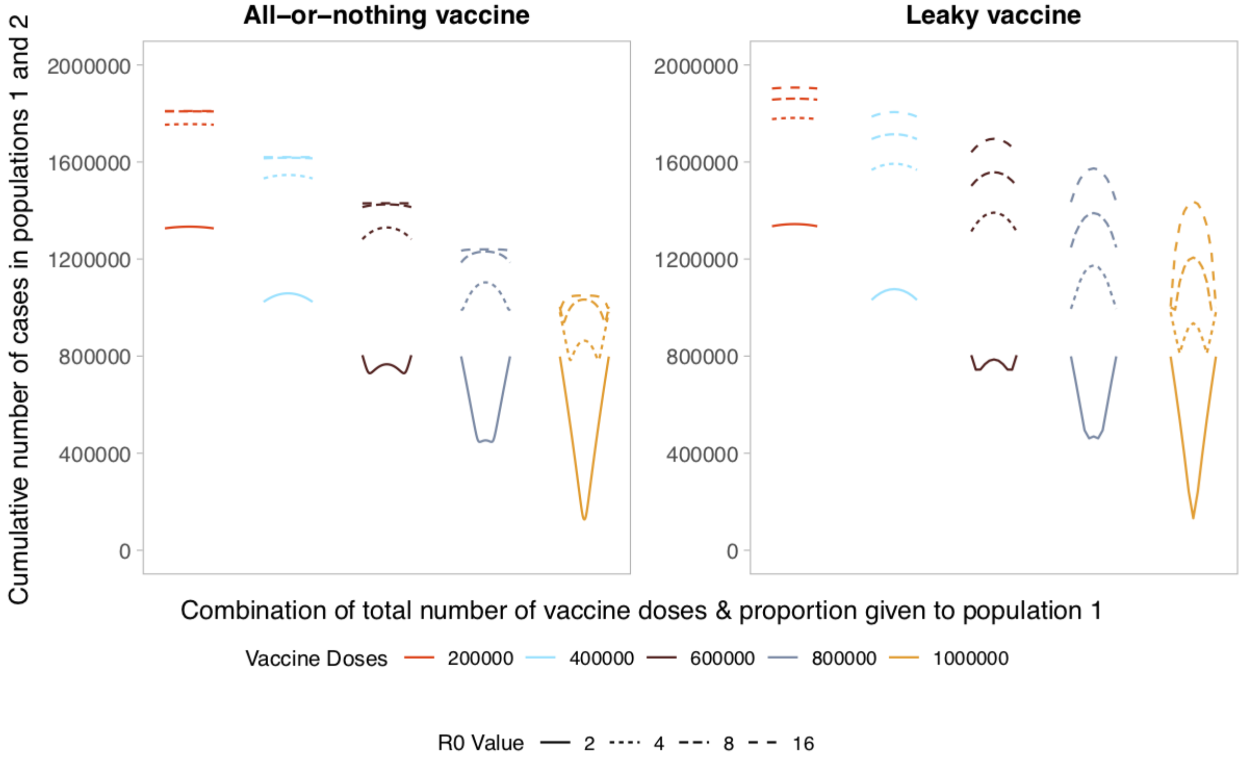


**Figure S1**: Performance of different allocation strategies of a limited vaccine stockpile across two homogeneous populations of equal size (one million individuals) with no underlying immunity and prophylactic vaccination. An all-or-nothing vaccine (left) is compared to a leaky vaccine (right). The panel on the left is equivalent to Figure 1.

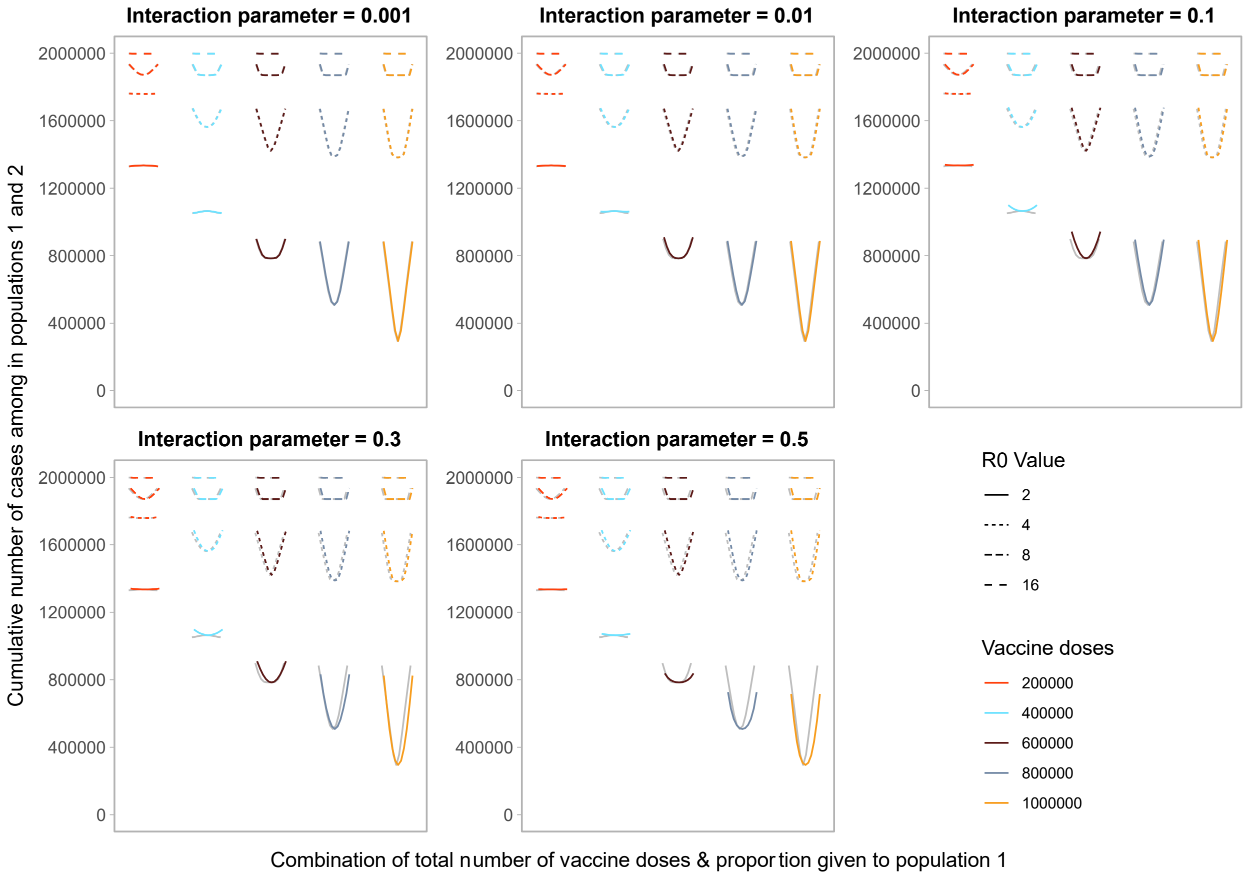


**Figure S2**: Performance of different allocation strategies of a limited vaccine stockpile across two homogeneous populations of equal size (one million individuals) with no underlying immunity, with vaccines rolled out starting 10 days after the start of the epidemic with 2% of the population vaccinated each day. The populations are allowed to interact of varying degree (i=0 corresponds no interaction and i=0.5 corresponds to complete interaction or perfect mixing between the two populations). The grey lines in each panel are equivalent to Figure 1 and represent a scenario without interaction between the two populations.

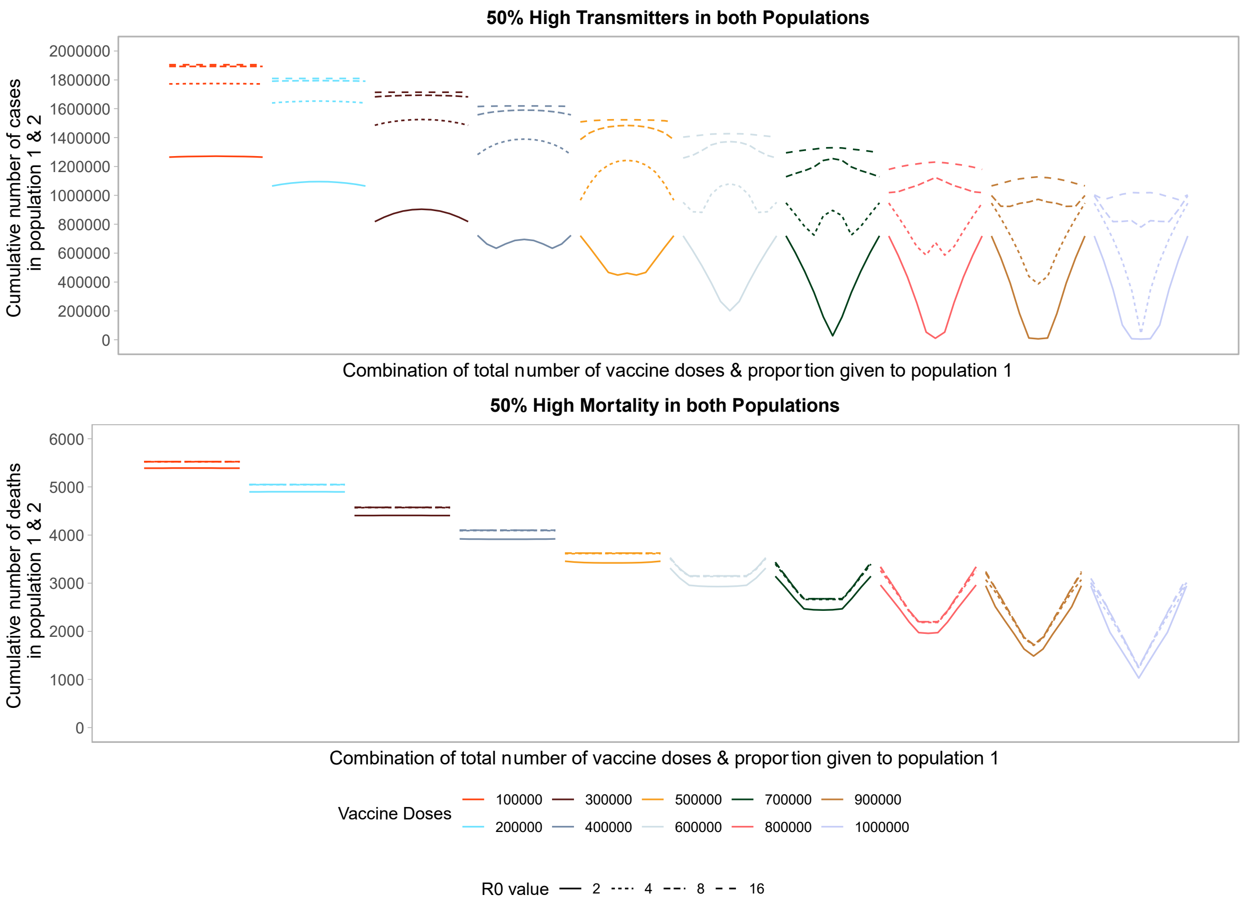


**Figure S3**: Performance of different allocation strategies of a limited vaccine stockpile across two heterogeneous populations of equal size (one million individuals) with no underlying immunity and prophylactic vaccination. Each color represents a different number of total vaccine doses. Each line represents a different basic reproductive number. In both the high transmission scenario (top) and high mortality scenario (bottom), 50% of both populations are high risk.

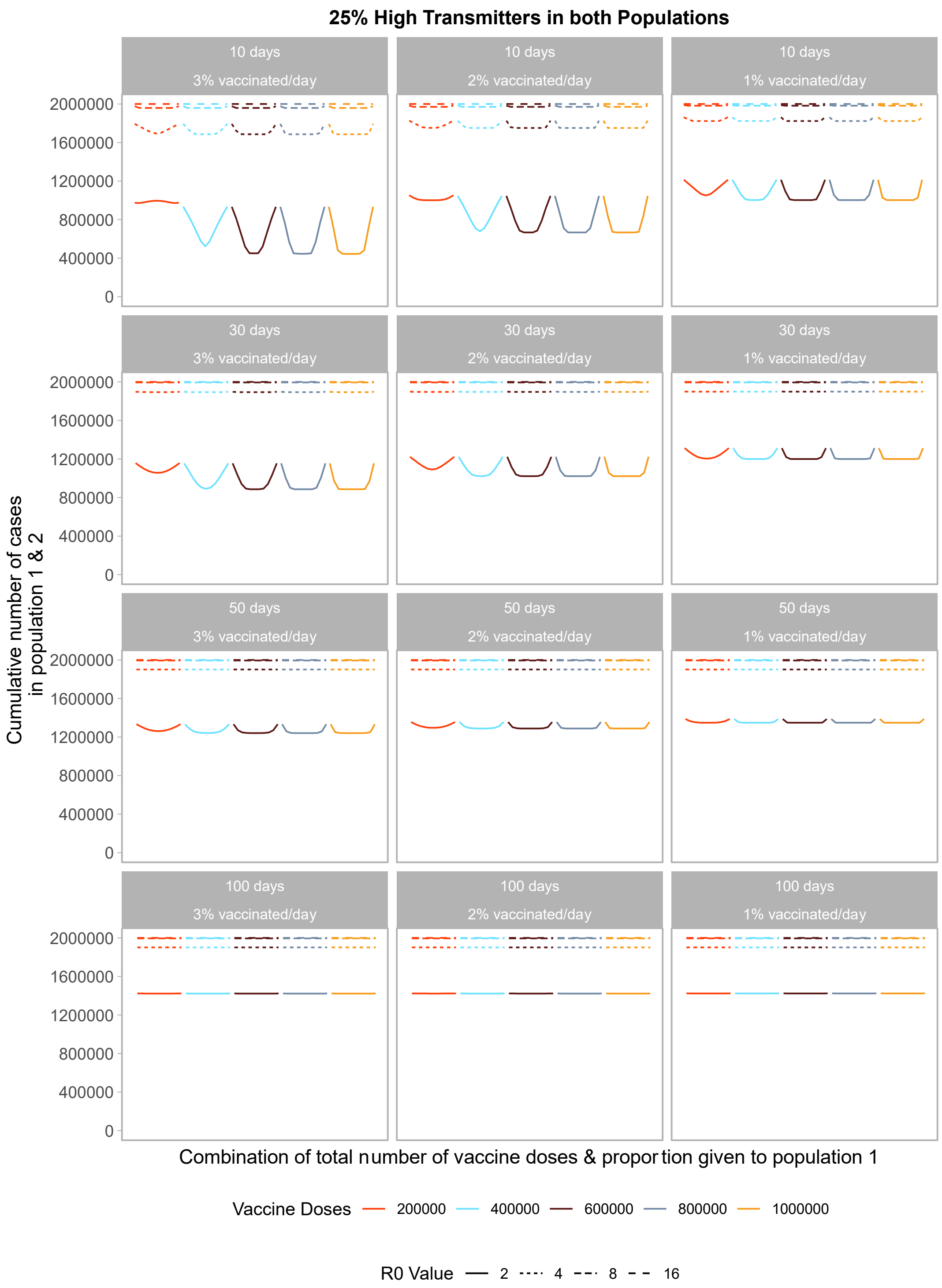


**Figure S4:** Performance of different allocation strategies of a limited vaccine stockpile across two heterogeneous populations of equal size (one million individuals) with no underlying immunity, with vaccines rolled out at different speeds and different times after the start of the epidemic. 25% of both populations are high risk of transmission. We vary the timing of roll-out between 10, 30, 50, or 100 days after the start of the epidemic, and vary the speed of roll-out between 1, 2, or 3% of the population vaccinated per day.

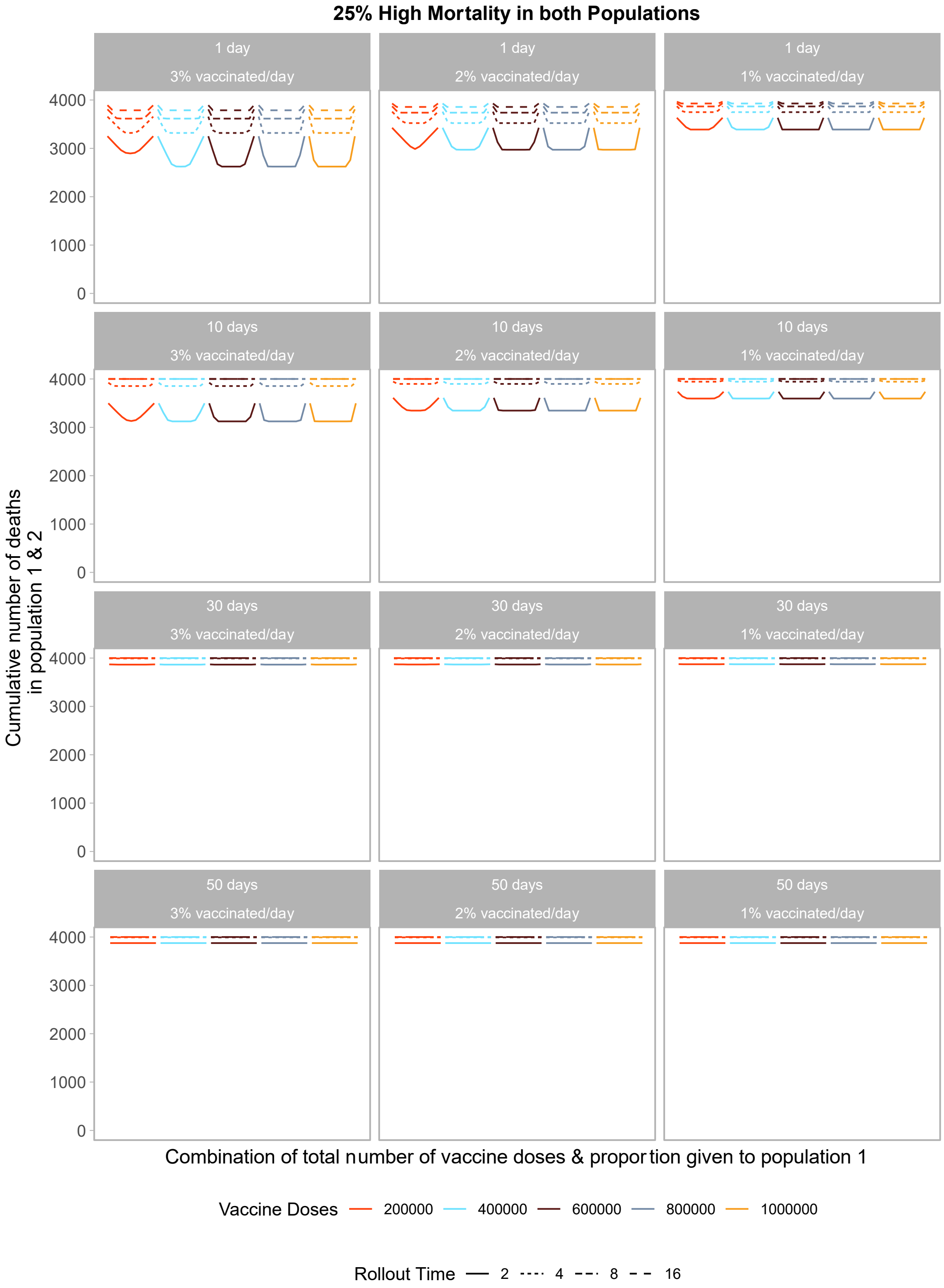


**Figure S5:** Performance of different allocation strategies of a limited vaccine stockpile across two heterogeneous populations of equal size (one million individuals) with no underlying immunity, with vaccines rolled out at different speeds and different times after the start of the epidemic. 25% of both populations are high risk of mortality. We vary the timing of roll-out between 1, 10, 30 or 50 days after the start of the epidemic and vary the speed of roll-out between 1, 2, or 3% of the population vaccinated per day.

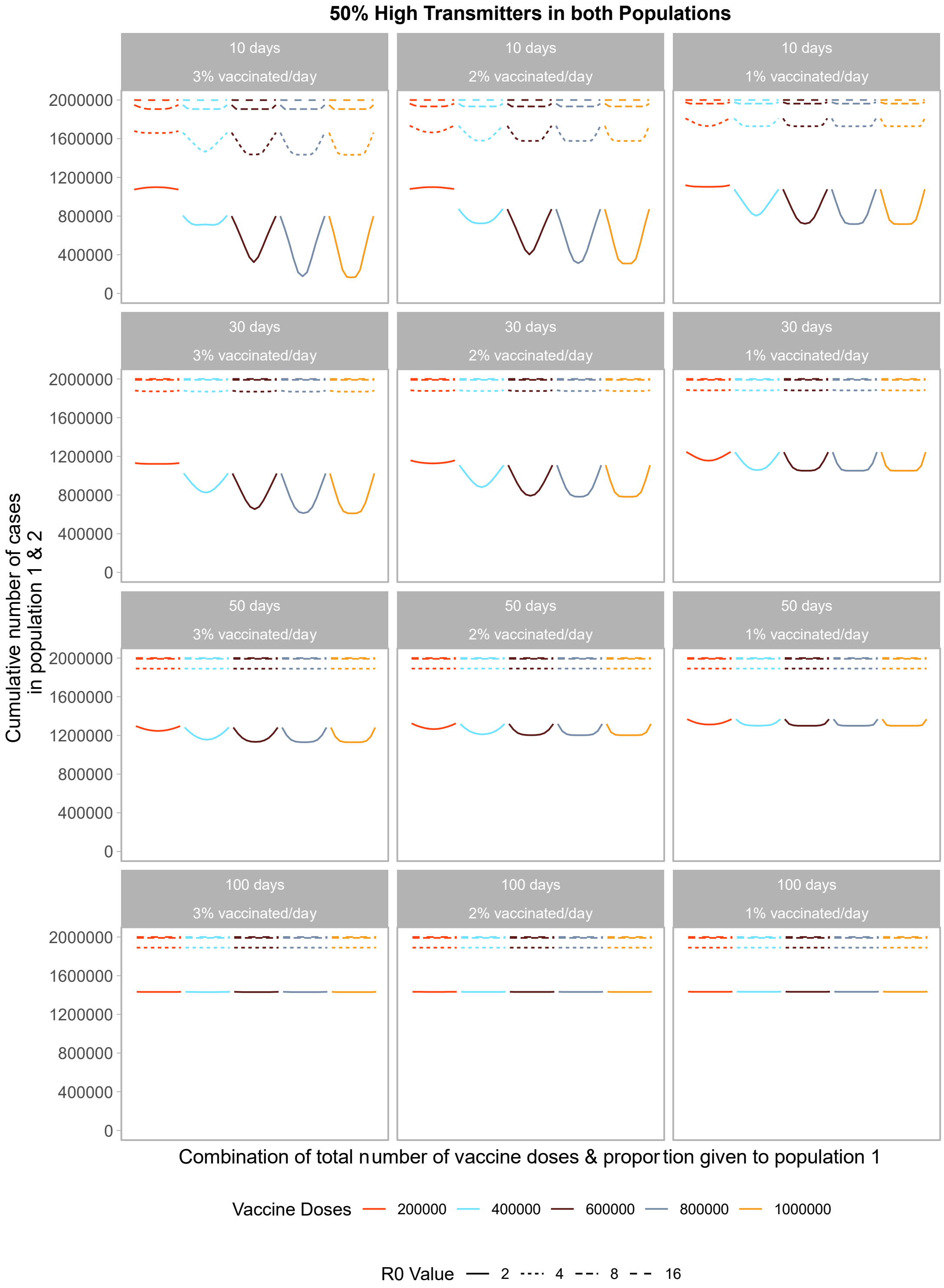


**Figure S6:** Performance of different allocation strategies of a limited vaccine stockpile across two heterogeneous populations of equal size (one million individuals) with no underlying immunity, with vaccines rolled out at different speeds and different times after the start of the epidemic. 50% of both populations are high risk of transmission. We vary the timing of roll-out between 10, 30, 50, or 100 days after the start of the epidemic, and vary the speed of roll-out between 1, 2, or 3% of the population vaccinated per day.

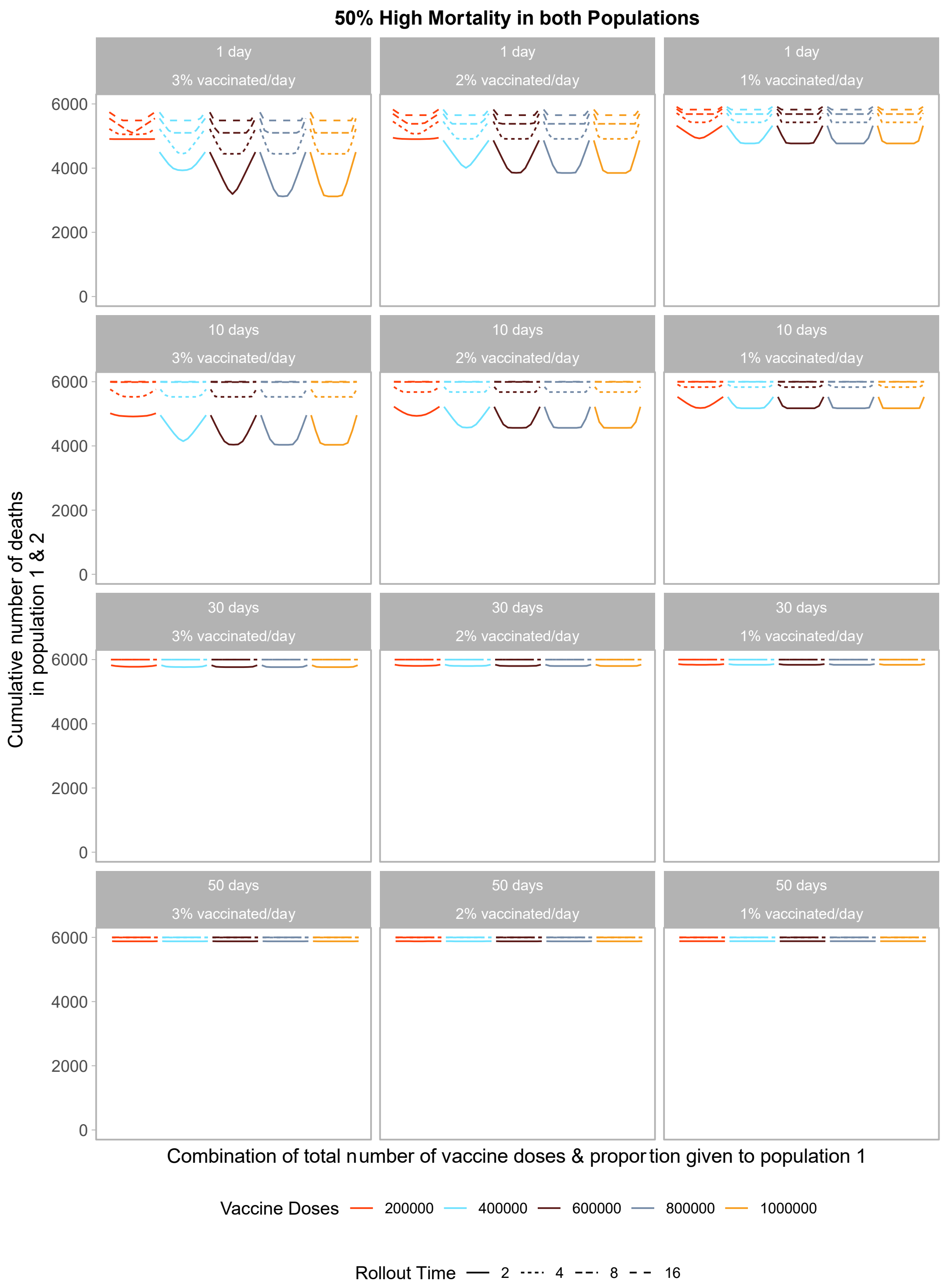


**Figure S7:** Performance of different allocation strategies of a limited vaccine stockpile across two heterogeneous populations of equal size (one million individuals) with no underlying immunity, with vaccines rolled out at different speeds and different times after the start of the epidemic. 50% of both populations are high risk of mortality. We vary the timing of roll-out between 1, 10, 30 or 50 days after the start of the epidemic and vary the speed of roll-out between 1, 2, or 3% of the population vaccinated per day.

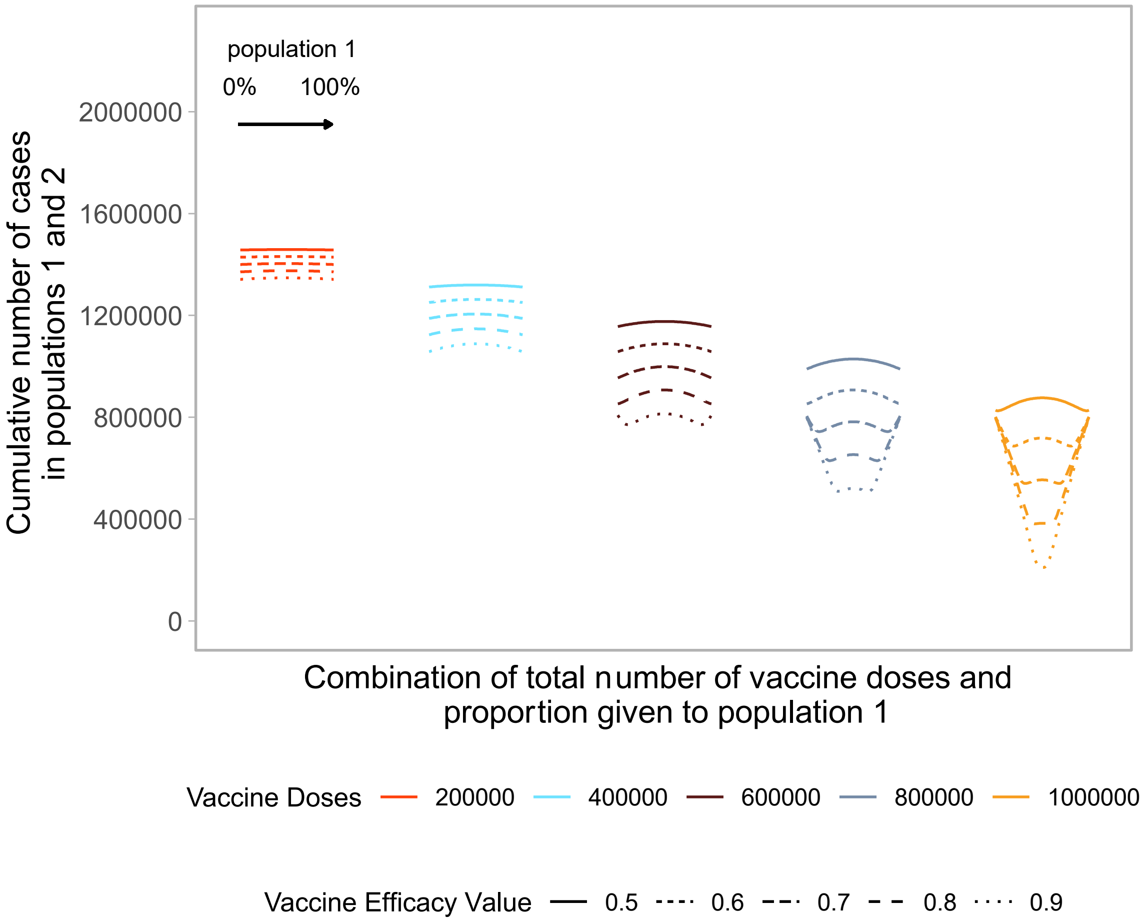


**Figure S8:** Performance of different allocation strategies of a limited vaccine stockpile across two homogeneous populations of equal size (one million individuals) with no underlying immunity, and prophylactic vaccination. Each color represents a different number of total vaccine doses. Each line represents a different vaccine efficacy value from 50 to 90%.

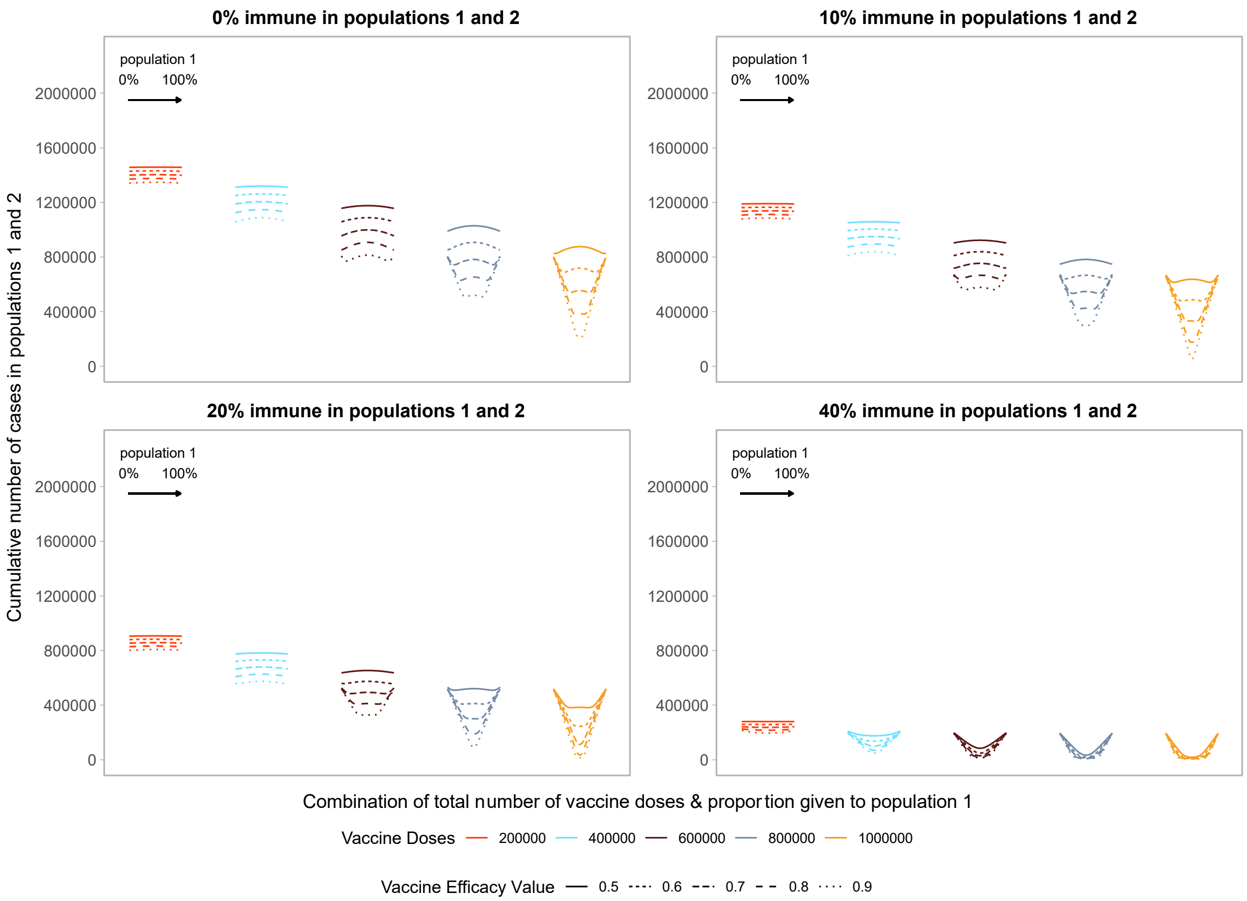


**Figure S9:** Performance of different allocation strategies of a limited vaccine stockpile across two homogeneous populations of equal size (one million individuals) with different underlying immunity, and prophylactic vaccination. We vary underlying immunity from 0 to 40%. The proportion if immune individuals is identical in both populations. Each color represents a different number of total vaccine doses. Each line represents a different vaccine efficacy value from 50 to 90%. The panel on the top left is equivalent to Figure S8.

#### A.2 Model and Parameters

##### A.2.1 SEIR model equations for two non-interacting populations

**Population 1**

$$\frac{dS_1}{dt} = -\beta S_1 I_1$$
$$\frac{dE_1}{dt} = \beta S_1 I_1 - \sigma E_1$$
$$\frac{dI_1}{dt} = \sigma E_1 - \nu I_1$$
$$\frac{dR_1}{dt} = \nu I_1$$

**Population 2**

$$\frac{dS_2}{dt} = -\beta S_2 I_2$$
$$\frac{dE_2}{dt} = \beta S_2 I_2 - \sigma E_2$$
$$\frac{dI_2}{dt} = \sigma E_2 - \nu I_2$$
$$\frac{dR_2}{dt} = \nu I_2$$

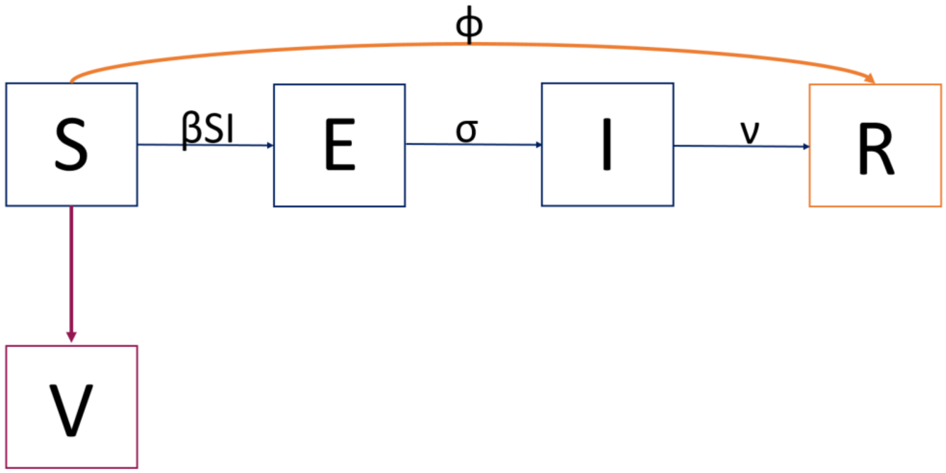


**Figure S8**: SEIR model incorporating underlying immunity (in orange) and continuous roll-out of vaccination (in purple).

##### A.2.2 Parameters

| Parameter | Definition | Value | Citation |
|---|---|---|---|
| $\sigma^{-1}$ | Latent period | 3 days | (Bubar et al., 2021; Kissler et al., 2020) |
| $\nu^{-1}$ | Infectious period | 5 days | (Bubar et al., 2021; Kissler et al., 2020) |
| $R_0$ | Basic reproduction number | 2, 4, 8, 16 | (Lee et al., 2010; McMorrow, 2021; Presanis et al., 2009) |
| $\tau$ | Vaccine efficacy | [0.5, 0.95] | (Doria-Rose et al., 2021; Tartof et al., 2021) |
| $v$ | Number of vaccine doses available | 1,000,000 | |
| $pv_1$ | Proportion of the total vaccines given to population 1 | [0,1] | |
| $\varphi_i$ | Proportion of the population immune in population 1 | 0, 0.1, 0.2, 0.4 | |
| Rollout time | Vaccine roll-out time | 1, 10, 30, 50, 100 days | |
| Rollout speed | Vaccine roll-out speed | 1, 2, 3% per day | |
| $ph_i$ | Proportion of high-risk individuals in population *i* | 0.25, 0.5 | |
| $i$ | Proportion of interaction in population 1 and 2 | [0,0.5] | |
| $s_h$ | Proportion of high-risk infected individuals that do not die in the model accounting for heterogeneous risk of mortality | 0.995 | (Williamson et al., 2020) |
| $s_l$ | Proportion of low-risk infected individuals that do not die in the model accounting for heterogeneous risk of mortality | 0.999 | (Williamson et al., 2020) |

##### A.2.3 SEIR model equations for two interacting populations

We extend the equations from the original SEIR model to allow for interaction between population 1 and population 2.

**Population 1**

$$\frac{dS_1}{dt} = -\beta S_1[(1-i)I_1 + iI_2]$$
$$\frac{dE_1}{dt} = \beta S_1[(1-i)I_1 + iI_2] - \sigma E_1$$
$$\frac{dI_1}{dt} = \sigma E_1 - \nu I_1$$
$$\frac{dR_1}{dt} = \nu I_1$$

**Population 2**

$$\frac{dS_2}{dt} = -\beta S_2[(1-i)I_2 + iI_1]$$
$$\frac{dE_2}{dt} = \beta S_2[(1-i)I_2 + iI_1] - \sigma E_2$$
$$\frac{dI_2}{dt} = \sigma E_2 - \nu I_2$$
$$\frac{dR_2}{dt} = \nu I_2$$

##### A.2.4 SEIR model equations for high-risk of transmission

We extend the equations from the original SEIR model to allow individuals at higher risk of transmission.

**Population 1**

$$\frac{dS_{1H}}{dt} = -\beta_{HH}S_{1H}I_{1H} - \beta_{HL}S_{1H}I_{1L}$$
$$\frac{dE_{1H}}{dt} = \beta_{HH}S_{1H}I_{1H} + \beta_{HL}S_{1H}I_{1L} - \sigma E_{1H}$$
$$\frac{dI_{1H}}{dt} = \sigma E_{1H} - \nu I_{1H}$$
$$\frac{dR_{1H}}{dt} = s_h \nu I_{1H}$$
$$\frac{dS_{1L}}{dt} = -\beta_{LL}S_{1L}I_{1L} - \beta_{LH}S_{1L}I_{1H}$$
$$\frac{dE_{1L}}{dt} = \beta_{LL}S_{1L}I_{1L} + \beta_{LH}S_{1L}I_{1H} - \sigma E_{1L}$$
$$\frac{dI_{1L}}{dt} = \sigma E_{1L} - \nu I_{1L}$$
$$\frac{dR_{1L}}{dt} = s_l \nu I_{1L}$$

**Population 2**

$$\frac{dS_{2H}}{dt} = -\beta_{HH}S_{2H}I_{2H} - \beta_{HL}S_{2H}I_{2L}$$

$$\frac{dE_{2H}}{dt} = \beta_{HH}S_{2H}I_{2H} + \beta_{HL}S_{2H}I_{2L} - \sigma E_{2H}$$
$$\frac{dI_{2H}}{dt} = \sigma E_{2H} - \nu I_{2H}$$
$$\frac{dR_{2H}}{dt} = s_h \nu I_{2H}$$
$$\frac{dS_{2L}}{dt} = -\beta_{LL}S_{2L}I_{2L} - \beta_{LH}S_{2L}I_{2H}$$
$$\frac{dE_{2L}}{dt} = \beta_{LL}S_{2L}I_{2L} + \beta_{LH}S_{2L}I_{2H} - \sigma E_{2L}$$
$$\frac{dI_{2L}}{dt} = \sigma E_{2L} - \nu I_{2L}$$
$$\frac{dR_{2L}}{dt} = s_l \nu I_{2L}$$

##### A.2.5 SEIR model equations for high-risk of mortality

We extend the equations from the original SEIR model to allow individuals at higher risk of death.

**Population 1**

$$\frac{dS_{1H}}{dt} = -\beta S_{1H}I_{1H} - \beta S_{1H}I_{1L}$$
$$\frac{dE_{1H}}{dt} = \beta S_{1H}I_{1H} + \beta S_{1H}I_{1L} - \sigma E_{1H}$$
$$\frac{dI_{1H}}{dt} = \sigma E_{1H} - \nu I_{1H}$$
$$\frac{dR_{1H}}{dt} = s_h \nu I_{1H}$$
$$\frac{dD_{1H}}{dt} = (1 - s_h)\nu I_{1H}$$

$$\frac{dS_{1L}}{dt} = -\beta S_{1L}I_{1L} - \beta S_{1L}I_{1H}$$
$$\frac{dE_{1L}}{dt} = \beta S_{1L}I_{1L} + \beta S_{1L}I_{1H} - \sigma E_{1L}$$
$$\frac{dI_{1L}}{dt} = \sigma E_{1L} - \nu I_{1L}$$
$$\frac{dR_{1L}}{dt} = s_l \nu I_{1L}$$
$$\frac{dD_{1L}}{dt} = (1 - s_l)\nu I_{1L}$$

**Population 2**

$$\frac{dS_{2H}}{dt} = -\beta S_{2H}I_{2H} - \beta S_{2H}I_{2L}$$
$$\frac{dE_{2H}}{dt} = \beta S_{2H}I_{2H} + \beta S_{2H}I_{2L} - \sigma E_{2H}$$
$$\frac{dI_{2H}}{dt} = \sigma E_{2H} - \nu I_{2H}$$

$$\frac{dR_{2H}}{dt} = s_h \nu I_{2H}$$
$$\frac{dD_{2H}}{dt} = (1 - s_h)\nu I_{2H}$$

$$\frac{dS_{2L}}{dt} = -\beta S_{2L} I_{2L} - \beta S_{2L} I_{2H}$$
$$\frac{dE_{2L}}{dt} = \beta S_{2L} I_{2L} + \beta S_{2L} I_{2H} - \sigma E_{2L}$$
$$\frac{dI_{2L}}{dt} = \sigma E_{2L} - \nu I_{2L}$$
$$\frac{dR_{2L}}{dt} = s_l \nu I_{2L}$$
$$\frac{dD_{2L}}{dt} = (1 - s_l)\nu I_{2L}$$

##### A.2.6 Global $R_0$ calculation for heterogeneous transmission

To identify the global basic reproduction number for the population in simulations with multiple types of individuals in the population, we use the next generation matrix for the SEIR model (van den Driessche, 2017), with two compartments for each of the SEIR components, one for the high-transmitters individuals and one for low-transmitter individuals. Letting $S_L^*$ and $S_H^*$ denote the disease-free-equilibrium proportion of individuals in the low-transmitters and high-transmitters susceptible compartments, respectively, and letting $\beta_{LL}$ be the force of transmission from one low-transmitter individual to another, $\beta_{LH}$ from a high-transmitter individual to a low-transmitter individual, $\beta_{HL}$ from a low-transmitter individual to a high-transmitter individual, and $\beta_{HH}$ from one high-transmitter individual to another, we get the components of the next generation matrix:

$$F = \begin{pmatrix} 0 & 0 & \beta_{LL} S_L^* & \beta_{LH} S_L^* \\ 0 & 0 & \beta_{HL} S_H^* & \beta_{HH} S_H^* \\ 0 & 0 & 0 & 0 \\ 0 & 0 & 0 & 0 \end{pmatrix}$$

$$V = \begin{pmatrix} \sigma & 0 & 0 & 0 \\ 0 & \sigma & 0 & 0 \\ -\sigma & 0 & \nu & 0 \\ 0 & -\sigma & 0 & \nu \end{pmatrix}$$

And thus:

$$V^{-1} = \begin{pmatrix} \sigma^{-1} & 0 & 0 & 0 \\ 0 & \sigma^{-1} & 0 & 0 \\ \nu^{-1} & 0 & \nu^{-1} & 0 \\ 0 & \nu^{-1} & 0 & \nu^{-1} \end{pmatrix}$$

$$FV^{-1} = \begin{pmatrix} \nu^{-1}\beta_{LL} S_L^* & \nu^{-1}\beta_{LH} S_L^* & \nu^{-1}\beta_{LL} S_L^* & \nu^{-1}\beta_{LH} S_L^* \\ \nu^{-1}\beta_{HL} S_H^* & \nu^{-1}\beta_{HH} S_H^* & \nu^{-1}\beta_{HL} S_H^* & \nu^{-1}\beta_{HH} S_H^* \\ 0 & 0 & 0 & 0 \\ 0 & 0 & 0 & 0 \end{pmatrix}$$

The spectral radius of $FV^{-1}$ is then given by the spectral radius of the upper left 2×2 submatrix:

$$\rho(FV^{-1}) = max\left\{|\lambda|: \begin{vmatrix} \nu^{-1}\beta_{LL}S_L^* - \lambda & \nu^{-1}\beta_{LH}S_L^* \\ \nu^{-1}\beta_{HL}S_H^* & \nu^{-1}\beta_{HH}S_H^* - \lambda \end{vmatrix} = 0\right\}$$
$$= max\{|\lambda|: (\beta_{LL}S_L^* - \lambda\nu)(\beta_{HH}S_H^* - \lambda\nu) - \beta_{LH}S_L^*\beta_{HL}S_H^* = 0\}$$
$$= max\{|\lambda|: (\lambda\nu)^2 - (\beta_{LL}S_L^* + \beta_{HH}S_H^*)(\lambda\nu) + \beta_{LL}S_L^*\beta_{HH}S_H^* - \beta_{LH}S_L^*\beta_{HL}S_H^* = 0\}$$
$$= max\left\{|\lambda|: \lambda\nu = \frac{\beta_{LL}S_L^* + \beta_{HH}S_H^*}{2} \pm \frac{\sqrt{(\beta_{LL}S_L^* + \beta_{HH}S_H^*)^2 - 4(\beta_{LL}S_L^* + \beta_{HH}S_H^* - \beta_{LH}S_L^*\beta_{HL}S_H^*)}}{2}\right\}$$
$$= \frac{\beta_{LL}S_L^* + \beta_{HH}S_H^* + \sqrt{(-\beta_{LL}S_L^* - \beta_{HH}S_H^*)^2 + 4\beta_{LH}S_L^*\beta_{HL}S_H^*}}{2\nu}$$
$$= \frac{R_{LL} + R_{HH} + \sqrt{(R_{LL} - R_{HH})^2 + 4R_{LH}R_{HL}}}{2}$$

This value is the global $R_0$.

$R_{HL}$ and $R_{LH}$ represent the number of secondary infections in high-transmitters generated by an infected low-transmitter and the number of secondary infections in a low-transmitter generated by an infected high-transmitter, respectively. $R_{HH}$ and $R_{LL}$ represent the number of secondary infections in high-transmitter members generated by an infected high-transmitter and the number of secondary infections in a low-transmitter generated by an infected low-transmitter.

###### A.2.7 Vaccine Allocation decision rules

Below we describe the decision rules when allocating a limited number of vaccines to two populations with heterogeneous risk structure.

1. Assign doses to either population 1 or population 2. This is determined by the *pv* parameter.

2. If the number of vaccine doses is greater than the population size ($v \times pv > N_1$) everyone in population 1 is vaccinated, and the leftover doses are assigned to the second population.

3. Once doses are assigned to each population, within each population we first assign all doses to the high-risk individuals and give any doses left to the low-risk individuals. This is accomplished by checking whether the number of doses are sufficient to cover all of the high-risk individuals in that population (i.e., $N_1 \times ph_1 < v \times pv_1$). If not, we assign all of the doses to high-risk individuals and none to low-risk individuals. If the number of doses are sufficient to cover all high-risk individuals, all of those individuals are vaccinated and the remaining doses are assigned to low-risk individuals.

#### A.3 Literature

###### A.3.1 Optimal allocation across populations papers

https://docs.google.com/spreadsheets/d/1NsWWBcztpGG4IpU2U65cUUMryNW9IoKQnoWyd6zig50/edit?usp=sharing

###### A.3.2 Details on optimal allocation threshold

Duijzer et al. (L. E. Duijzer et al., 2018) identify many features of the direct and indirect effect of vaccination that determine the optimal threshold to which to vaccinate populations. The authors seek to minimize the final size of the epidemic or, equivalently, maximize the total number of people who escape infection. When the number of vaccine doses available is less than the herd immunity threshold, this is also equivalent to maximizing the number of susceptible individuals remaining at the end of the epidemic, i.e., the number of unvaccinated individuals who escape infection, denoted the "herd effect." They determine that the herd effect, as a function of the vaccination fraction $f$ within a population, denoted $G(f)$, has a predictable structure: it is increasing and convex for a low value of $f$, until $\bar{f}$. From $\bar{f}$ to $f^*$, it is increasing and concave. Above $f^*$, $G$ is a decreasing function. $f^*$ is equivalent to the herd immunity threshold, equal to $1 - 1/R_0$ in a fully susceptible population. Vaccinating beyond $f^*$ decreases the herd effect, as individuals who would be (somewhat) protected through herd immunity are instead vaccinated and protected directly instead. This convex-concave structure occurs because, for low values of $f$, the epidemic peak is delayed in addition to being smaller in magnitude. Whereas for values of $f$ closer to $f^*$, the epidemic peak is advanced and smaller in magnitude. As $f$ increases to $f^*$, this earlier peak continues to advance, leading to a decline in the increase in the herd effect and a concave $G$ function. The authors identify another quantity: the dose-optimal vaccination fraction, $\tilde{f}$, where $\bar{f} \leq \tilde{f} \leq f^*$. The dose-optimal vaccination fraction maximizes the increase in herd effect per dose of vaccine. They find that for multiple non-interacting populations, the optimal allocation is to vaccinate as many populations as possible to the level $\tilde{f}$, and not vaccinate any other populations (except for perhaps one with any extra doses).

However, Duijzer et al. (L. E. Duijzer et al., 2018) prove that these features do not hold when there are no active infections (e.g., prior to the outbreak). In this case, $\bar{f} = \tilde{f} = f^*$. That is, $G$ is an increasing and convex function prior to the herd immunity threshold and a decreasing and concave function after the herd immunity threshold. In this case, the results of Duijzer et al. align with those of Keeling and Shattock (Keeling & Shattock, 2012): the optimal allocation scheme is to vaccinate as many populations as possible up to the herd immunity threshold. The difference from the previously described situation is that the peak is always delayed by increasing the vaccinations in a population with no active infections. Since pre-outbreak vaccination leads to a decreased transmission rate for all cases in the population, the reduced number needed to reach the pandemic peak is always outweighed by the increased time required to infect those individuals. Duijzer et al. (E. Duijzer et al., 2016) previously demonstrated that, in this case, maximizing the herd effect is achieved when $R_t = 1$, i.e. at the herd immunity threshold, so this comports with that finding as well.

### References:


Bubar, K. M., Reinholt, K., Kissler, S. M., Lipsitch, M., Cobey, S., Grad, Y. H., & Larremore, D. B. (2021). Model-informed COVID-19 vaccine prioritization strategies by age and serostatus. *Science*, *371*(6532), 916–921. https://doi.org/10.1126/science.abe6959

Doria-Rose, N., Suthar, M. S., Makowski, M., O'Connell, S., McDermott, A. B., Flach, B., Ledgerwood, J. E., Mascola, J. R., Graham, B. S., Lin, B. C., O'Dell, S., Schmidt, S. D., Widge, A. T., Edara, V.-V., Anderson, E. J., Lai, L., Floyd, K., Rouphael, N. G., Zarnitsyna, V., … Kunwar, P. (2021). Antibody Persistence through 6 Months after the Second Dose of mRNA-1273 Vaccine for Covid-19. *New England Journal of Medicine*, *384*(23), 2259–2261. https://doi.org/10.1056/NEJMc2103916

Duijzer, E., van Jaarsveld, W., Wallinga, J., & Dekker, R. (2016). The most efficient critical vaccination coverage and its equivalence with maximizing the herd effect. *Mathematical Biosciences*, *282*, 68–81. https://doi.org/10.1016/j.mbs.2016.09.017

Duijzer, L. E., van Jaarsveld, W. L., Wallinga, J., & Dekker, R. (2018). Dose-Optimal Vaccine Allocation over Multiple Populations. *Production and Operations Management*, *27*(1), 143–159. https://doi.org/10.1111/poms.12788

Keeling, M. J., & Shattock, A. (2012). Optimal but unequitable prophylactic distribution of vaccine. *Epidemics*, *4*(2), 78–85. https://doi.org/10.1016/j.epidem.2012.03.001

Kissler, S. M., Tedijanto, C., Goldstein, E., Grad, Y. H., & Lipsitch, M. (2020). Projecting the transmission dynamics of SARS-CoV-2 through the postpandemic period. *Science*, *368*(6493), 860–868. https://doi.org/10.1126/science.abb5793

Lee, B. Y., Brown, S. T., Korch, G. W., Cooley, P. C., Zimmerman, R. K., Wheaton, W. D., Zimmer, S. M., Grefenstette, J. J., Bailey, R. R., Assi, T.-M., & Burke, D. S. (2010). A computer simulation of vaccine prioritization, allocation, and rationing during the 2009 H1N1 influenza pandemic. *Vaccine*, *28*(31), 4875–4879. https://doi.org/10.1016/j.vaccine.2010.05.002

McMorrow, M. (2021). *Improving communications around vaccine breakthrough and vaccine effectiveness*. https://context-cdn.washingtonpost.com/notes/prod/default/documents/8a726408-07bd-46bd-a945-3af0ae2f3c37/note/57c98604-3b54-44f0-8b44-b148d8f75165

Presanis, A. M., De Angelis, D., Team3¶, T. N. Y. C. S. F. I., Hagy, A., Reed, C., Riley, S., Cooper, B. S., Finelli, L., Biedrzycki, P., & Lipsitch, M. (2009). The Severity of Pandemic H1N1 Influenza in the United States, from April to July 2009: A Bayesian Analysis. *PLOS Medicine*, *6*(12), e1000207. https://doi.org/10.1371/journal.pmed.1000207

Tartof, S. Y., Slezak, J. M., Fischer, H., Hong, V., Ackerson, B. K., Ranasinghe, O. N., Frankland, T. B., Ogun, O. A., Zamparo, J. M., Gray, S., Valluri, S. R., Pan, K., Angulo, F. J., Jodar, L., & McLaughlin, J. M. (2021). Effectiveness of mRNA BNT162b2 COVID-19 vaccine up to 6 months in a large integrated health system in the USA: a retrospective cohort study. *The Lancet*, *398*(10309), 1407–1416. https://doi.org/10.1016/S0140-6736(21)02183-8

van den Driessche, P. (2017). Reproduction numbers of infectious disease models. *Infectious Disease Modelling*, *2*(3), 288–303. https://doi.org/10.1016/j.idm.2017.06.002

Williamson, E. J., Walker, A. J., Bhaskaran, K., Bacon, S., Bates, C., Morton, C. E., Curtis, H. J., Mehrkar,

A., Evans, D., Inglesby, P., Cockburn, J., McDonald, H. I., MacKenna, B., Tomlinson, L., Douglas, I. J., Rentsch, C. T., Mathur, R., Wong, A. Y. S., Grieve, R., … Goldacre, B. (2020). Factors associated with COVID-19-related death using OpenSAFELY. *Nature*, *584*(7821), 430–436. https://doi.org/10.1038/s41586-020-2521-4
